# HIV and Pregnancy among Adolescents in Sub-Saharan Africa: A Scoping Review

**DOI:** 10.1101/2024.04.25.24305581

**Authors:** Folahanmi Tomiwa Akinsolu, Ifeoluwa Eunice Adewole, Abisola Lawale, Mobolaji Timothy Olagunju, Olunike Rebecca Abodunrin, Oluwabukola Mary Ola, Abel Nnamdi Chukwuemeka, Aisha Oluwaseun Gambari, George Uchenna Eleje, Oliver Chukwujekwu Ezechi

## Abstract

The interaction of adolescent pregnancy with HIV is a complex and crucial public health issue worldwide. This is evident in sub-Saharan Africa, where the co-occurrence of HIV and adolescent pregnancies represents a high risk for additional health and social consequences for both adolescent mothers and their children. This study aims to explore literature to provide a clear synthesis of data related to pregnancies among adolescents living with HIV in Sub-Saharan Africa.

A literature search was conducted in July 2023 using PubMed, Scopus, CINAHL, Cochrane Library, African Journals Online, and World Health Organization Global Index Medicus. Two authors screened potential studies. Articles selected were those published in or translated into the English language between 2002 and 2022.

Among 1,560 identified results, 25 peer-reviewed publications on pregnancy among adolescents living with HIV in Sub-Saharan Africa were included after screening. The studies, included both qualitative and quantitative data, with a total of 178,227 participants in the age range of 12-18 years. The majority of the studies reported quantitative data, while a few presented qualitative or mixed-methods findings.

This scoping review highlights disparities in pregnancy prevalence, revealing varied rates among different age groups. The underrepresentation of adolescents in demographic studies suggests diverse vulnerabilities and access to healthcare. The study also emphasizes the need for tailored healthcare services and comprehensive interventions addressing HIV risk, mental health challenges, and socio-economic barriers.

## INTRODUCTION

Global estimates show that every year, approximately 21 million pregnancies and 12 million births occur among adolescents in low- and middle-income countries [1]. Sub-Saharan Africa is responsible for some of the highest rates of adolescent pregnancies globally [2], with almost one in five females experiencing pregnancy between the ages of 10 and 19 [3].

Adolescent parents and pregnant adolescents face the challenge of managing pregnancy, parenting, and child-rearing responsibilities amid a comprehensive developmental phase often characterized by substantial psychological, social, and biological changes [4].

Furthermore, adolescents make up a large proportion of the people living with HIV globally (1.18 million-2.19 million). In 2022, 140,000 adolescents aged 10 to 19 were newly infected with HIV, with girls accounting for about 80% of them [5]. Particularly in Sub-Saharan Africa, nearly six times the number of newly acquired HIV infections occur among adolescent girls.

Adolescents are considerably more vulnerable to HIV infection compared to adults, given their ongoing developmental stage and the necessity to adapt to rapid biological, physical, and structural changes in their lives [5, 6]. The interaction of adolescent pregnancy with HIV is a complex and crucial public health issue worldwide. This is evident in sub-Saharan Africa, where the co-occurrence of HIV and adolescent pregnancies represents a high risk for additional health and social consequences for both adolescent mothers and their children [7].

With the high rates of early motherhood and HIV incidence in sub-Saharan Africa, designing targeted interventions, enhancing reproductive health services, and encouraging improved overall health outcomes are largely hinged on having this clear understanding of pregnancies among adolescents living with HIV in Sub-Saharan Africa. Thus, this scoping review aims to explore existing literature to provide a clear synthesis of data related to pregnancies among adolescents living with HIV in Sub-Saharan Africa.

## MATERIALS AND METHODS

### Overview

This review was guided by the methodological framework of Arksey and O’Malley [8]. Thus, the five steps followed were: (i) identifying the research question, (ii) identifying relevant studies, (iii) selecting eligible studies, (iv) charting the data, and (v) collating and summarizing the results. Appraisal of data quality was not done as the intention was to map all research activities in the field. The preferred reporting items for systematic reviews and meta-analyses extension for scoping reviews (PRISMA-ScR) checklist [9] was utilized with mapping done using the PRISMA-P chart [10].

We conducted a scoping review to synthesize all available evidence on pregnancies among adolescents aged 10 to 19 living with HIV in various Sub-Saharan African countries. A comprehensive search was conducted across major electronic databases, including PubMed, Scopus, CINAHL, Web of Science, and the World Health Organization Global Index Medicus, for studies published from 2002 up to December 2022. The scoping review adhered to the Preferred Reporting Items for Systematic Reviews and Meta-Analyses Extension for Scoping Reviews (PRISMA-ScR) guidelines [10] to ensure methodological rigour and transparency.

Articles published in English and reporting original data on pregnancies among adolescents living with HIV in Sub-Saharan Africa were included. Study selection and data extraction were conducted independently by four reviewers (O.O, O.A, A.L and I.A) to ensure the reliability of the findings. Conflicts were resolved by a fifth reviewer (F.T).

### Articles Identification

The initial search was conducted in July 2023 across major electronic databases, including PubMed, Scopus, Google Scholar, and the World Health Organization Global Index Medicus, for studies published from 2002 up to December 2022. The search was performed using the following search strategies enumerated in Appendix 1, combining keywords and Medical Subject Heading (MeSH) terms: “pregnancy”, “adolescents”, “HIV”, and “Sub-Saharan Africa”. The search strategy was refined to further check the appropriateness of the keywords and was expanded to include “females”, “prevalence”, “SSA”, “teenage pregnancy”, and “teenager”. A hand search was also conducted on the references of the included studies and websites of various postgraduate and undergraduate medical colleges to identify potentially relevant literature. The search and screening details are summarized with a graphical presentation using the PRISMA flow diagram in Figure 1. No protocol was published for this review.

The identification and analysis of data in this review was based on the Population/Exposure/Outcome (PEO) framework [11]. This framework was used to structure the review process by identifying key components of the main research question, which are population, exposure and outcome. In this review, the population of interest were adolescents living with HIV in Sub-Saharan Africa, the exposure is HIV status among adolescents in Africa, and the outcome is the prevalence of pregnancy among adolescents living with HIV in Africa.

### Search strategy

To achieve our study objectives, a comprehensive search was conducted across major electronic databases, including PubMed, Scopus, Google Scholar, and the World Health Organization Global Index Medicus, for studies published from 2002 up to December 2022.

In this scoping review, we searched for studies that reported pregnancy among female adolescents living with HIV in Sub-Saharan Africa. The search was conducted on studies published between 2002 and 2022 to obtain recent data on pregnancies among female adolescents living with HIV in Sub-Saharan Africa and its associated risk factors (Appendix 1).

### Eligibility Criteria and Article Selection

The literature obtained through database searches was imported into the reference management website, Rayyan.ai. In Rayyan, duplicates were removed using the “duplicates items” function. Two independent reviewers (M. O and I.A) conducted title and abstract screening, following the eligibility criteria set for this review, according to the PEO framework [11]. Full-text review of the remaining publications was then completed independently by four researchers (O.O, O.A, A.L and I.A) with reasons for inclusion or exclusion noted. Conflicts were resolved by F.T. Inclusion criteria required that the publications be in English and have full texts available to extract all relevant information. Studies were excluded if they were grey literature, theses, abstracts only or published before January 2002. Studies published between January 2002 and December 2022 were considered for inclusion. The titles and abstracts were used to screen the articles. Studies that do not report the following were excluded: pregnancy in adolescents living with HIV in a Sub-Saharan African Country; HIV status; sample size; lack primary data or well-defined methodology; duplicated samples and case-control studies and clinical trials; participants not adolescents; non-Sub-Saharan African women of known HIV status.

### Data Charting

From the publications included in this review, information on the paper identifiers (title, author, link), the country, year of publication, study aim, study design, study location, HIV status, adolescent pregnancy, and study findings were extracted. The extracted information from each publication was compiled and summarized in Table 1.

**Table 1:**
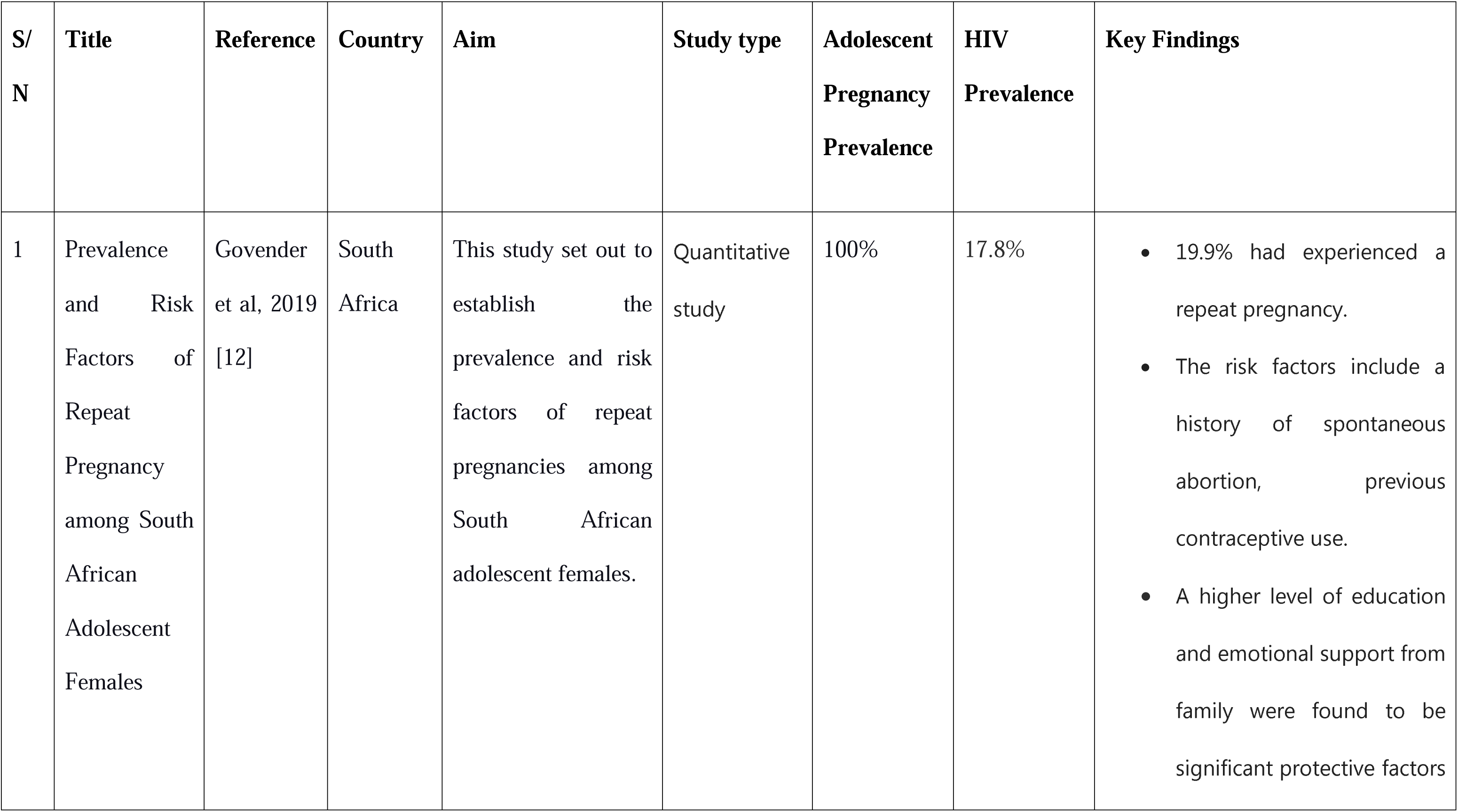
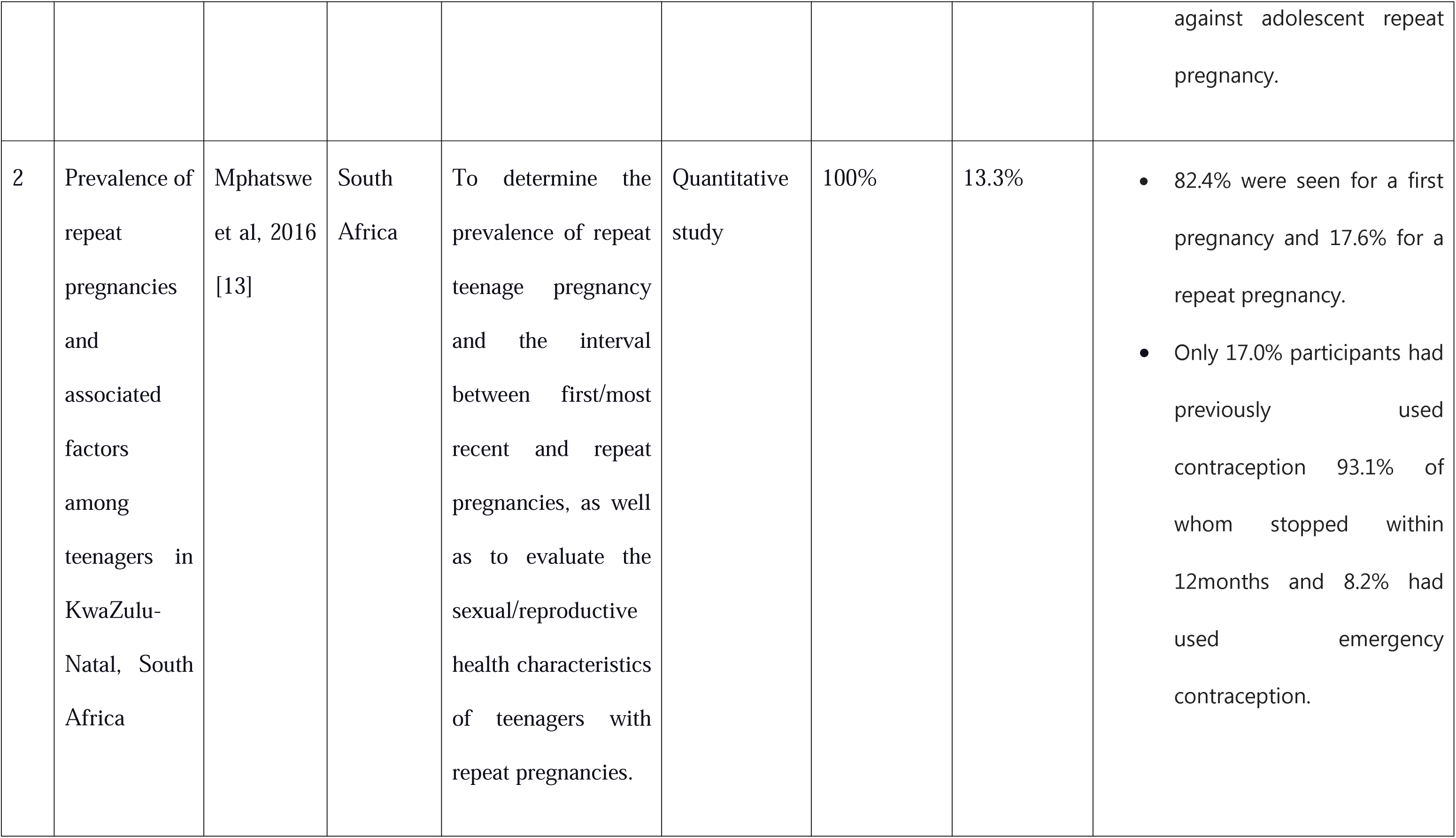
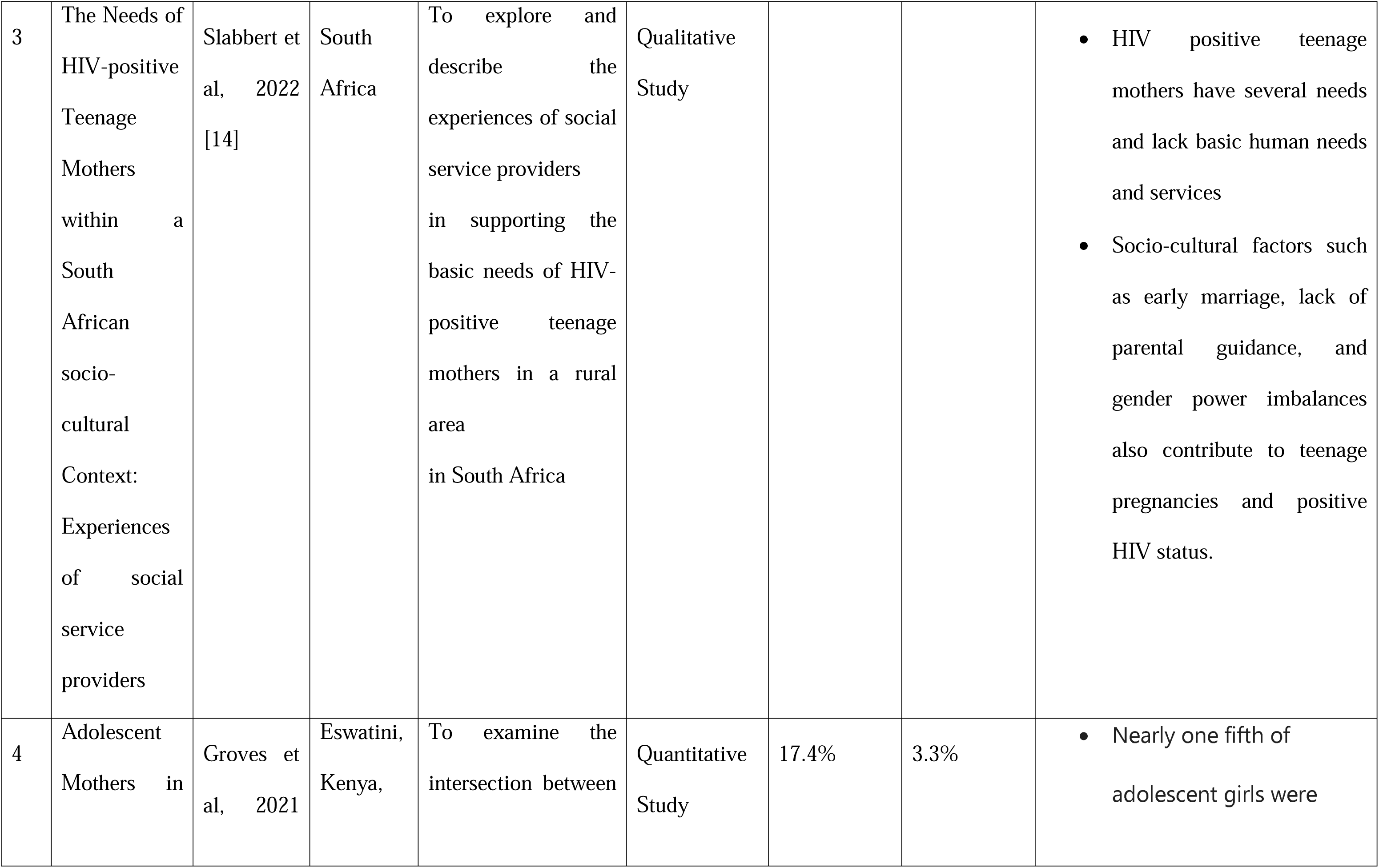
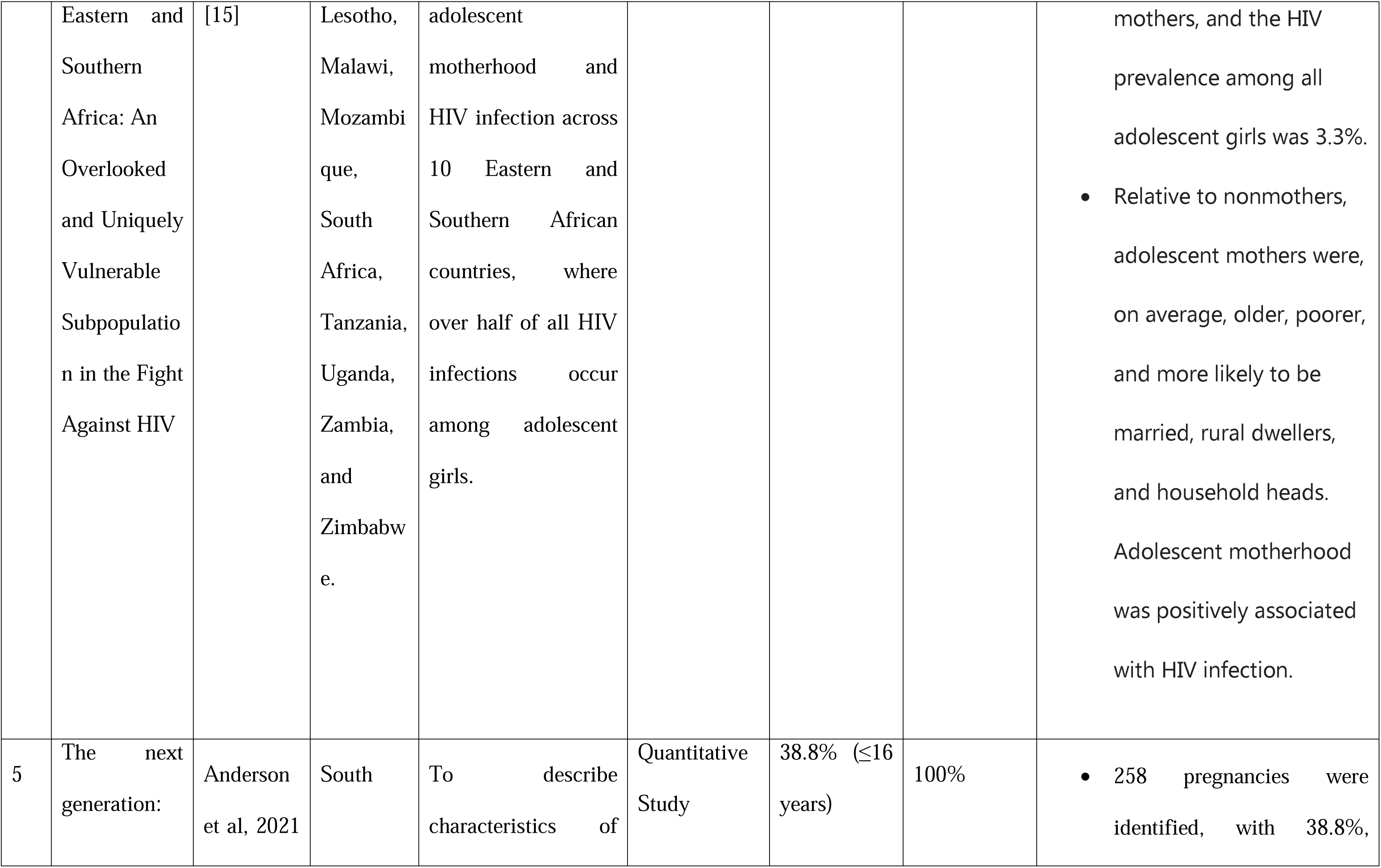
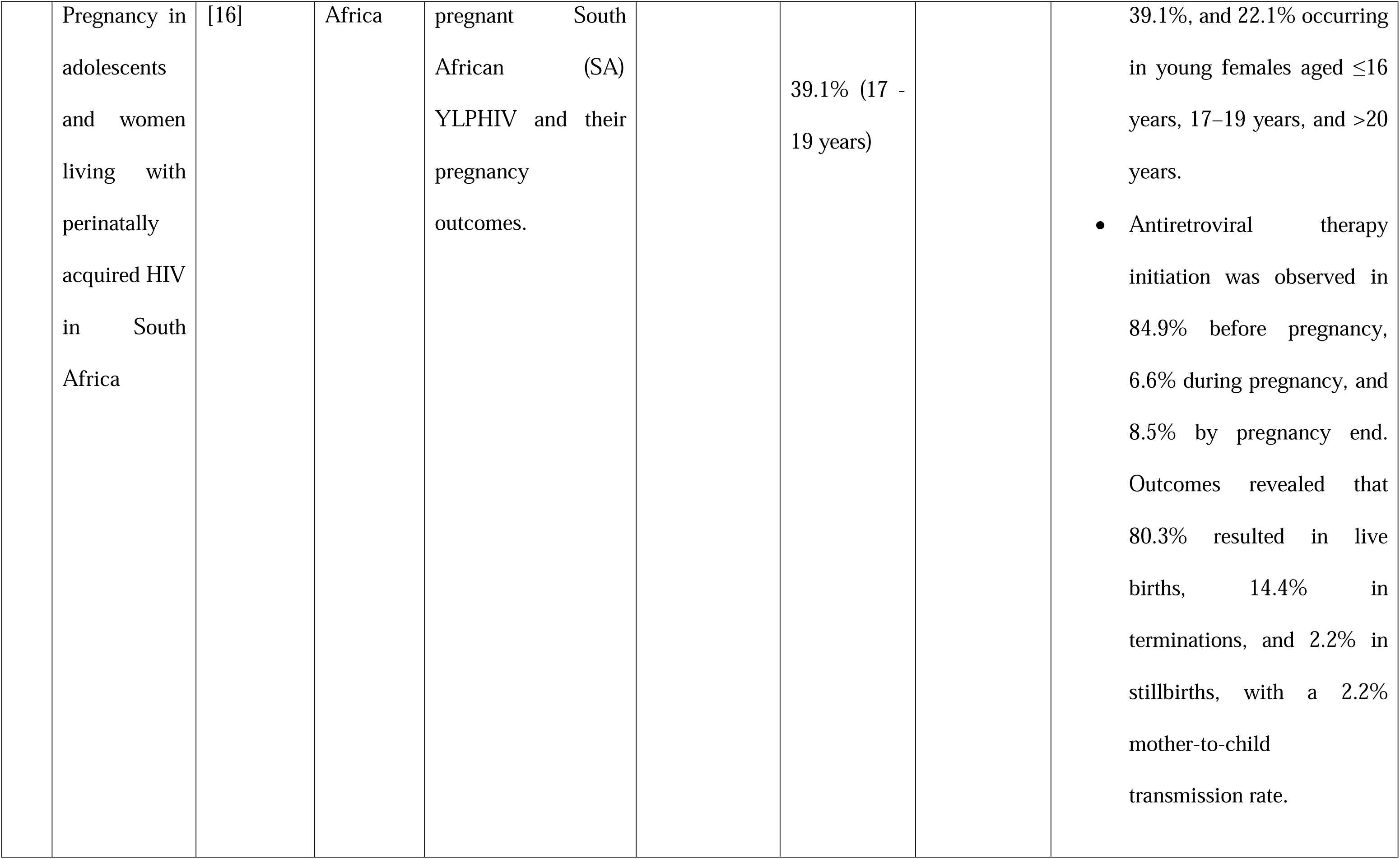
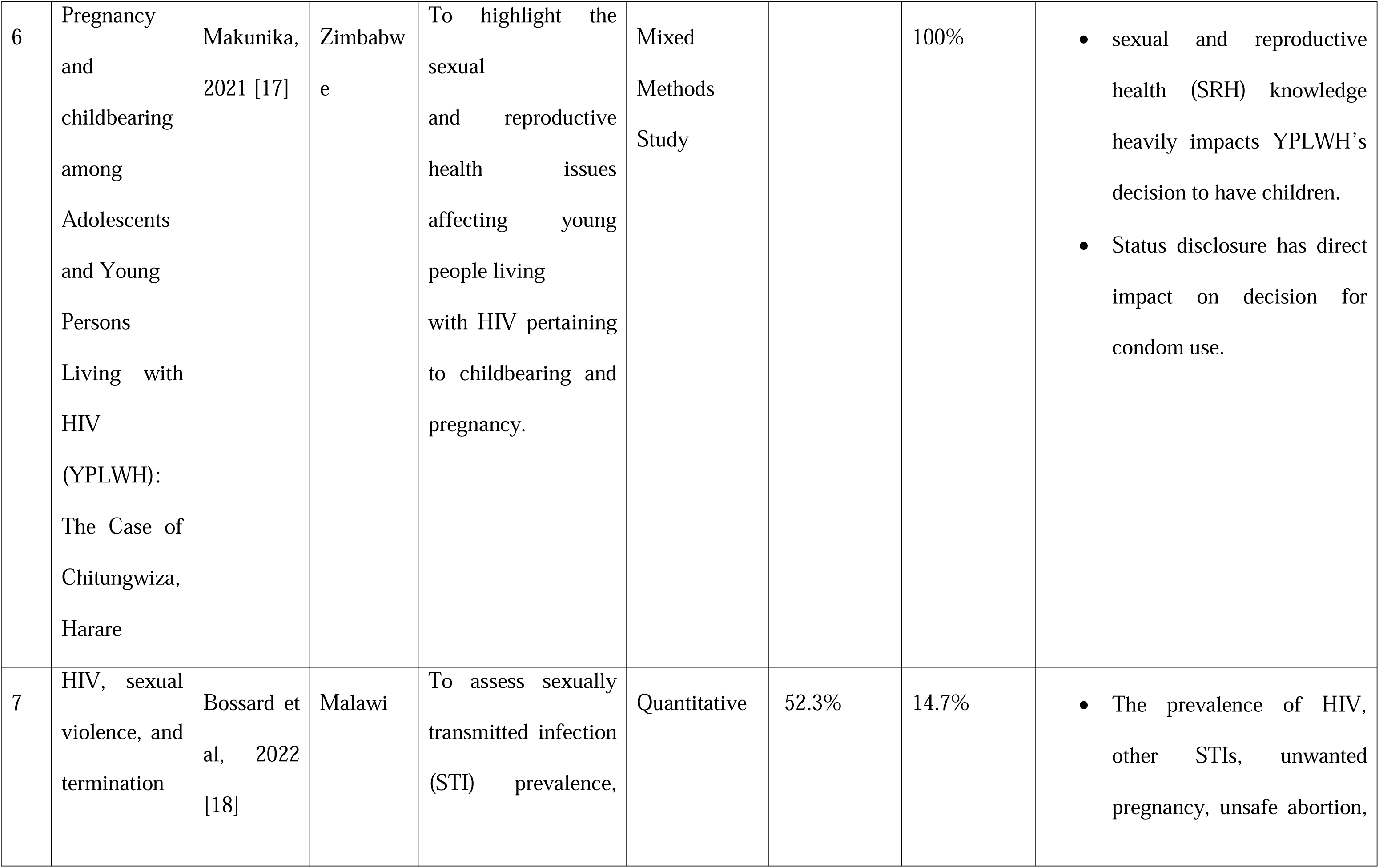
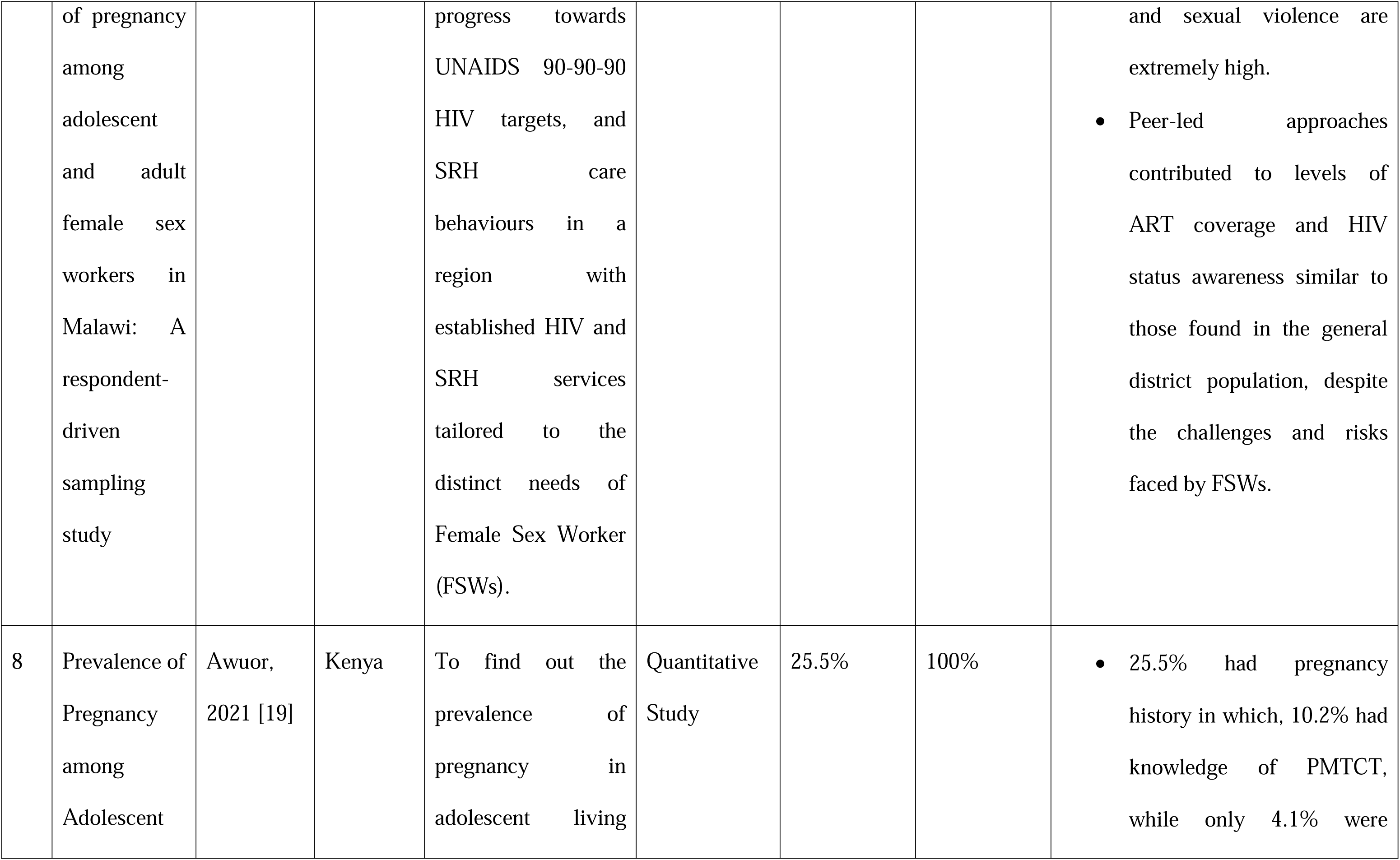
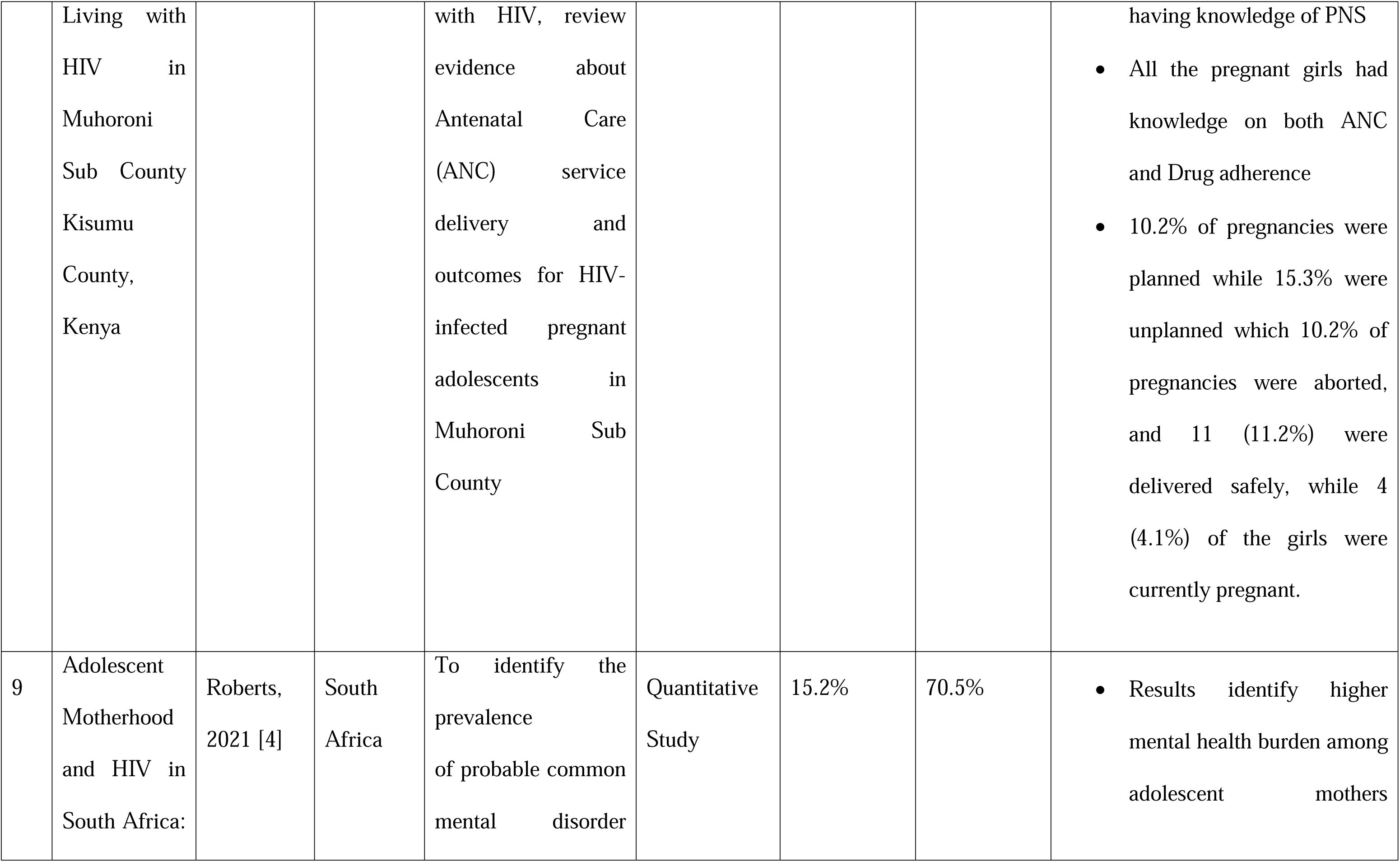
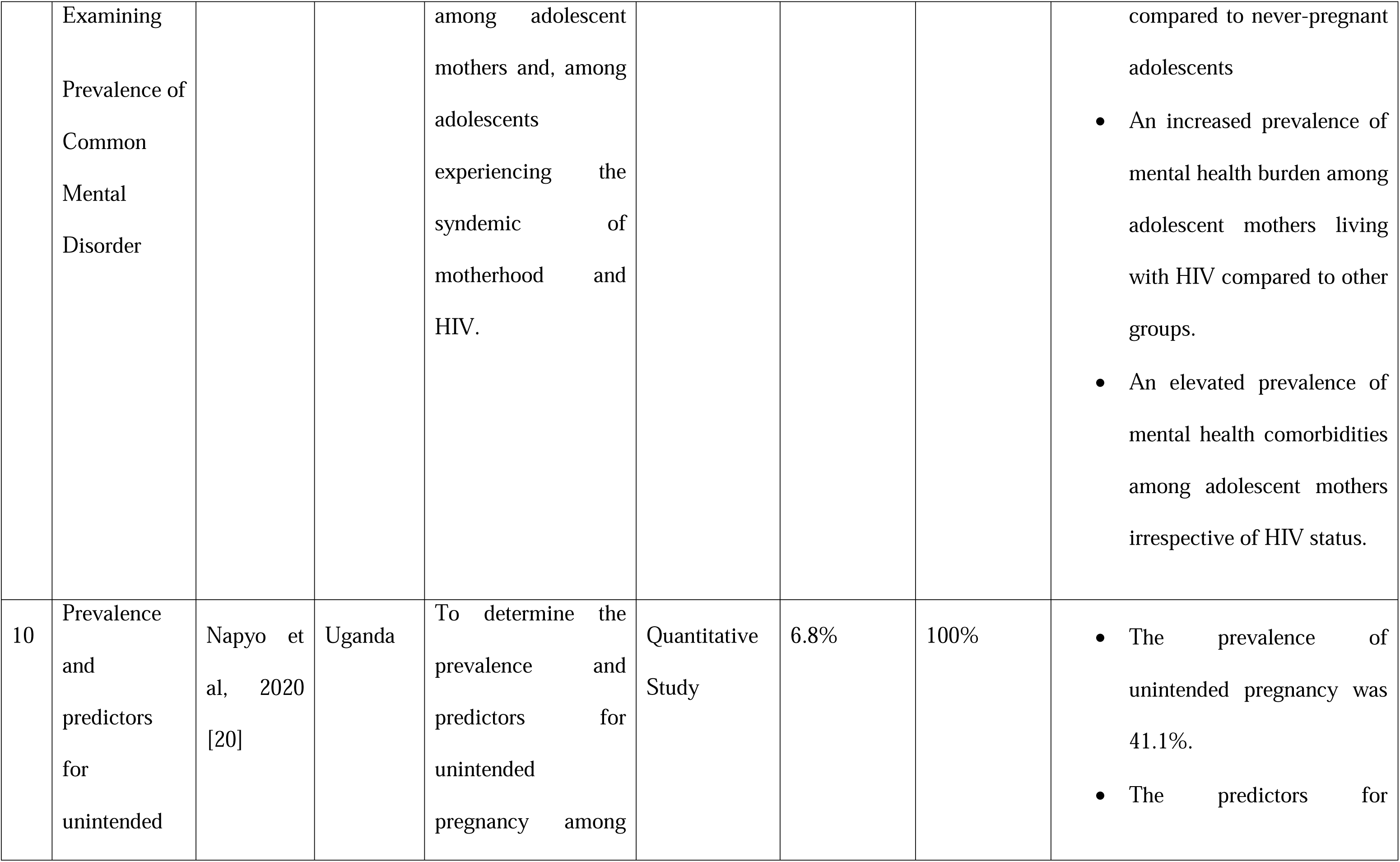
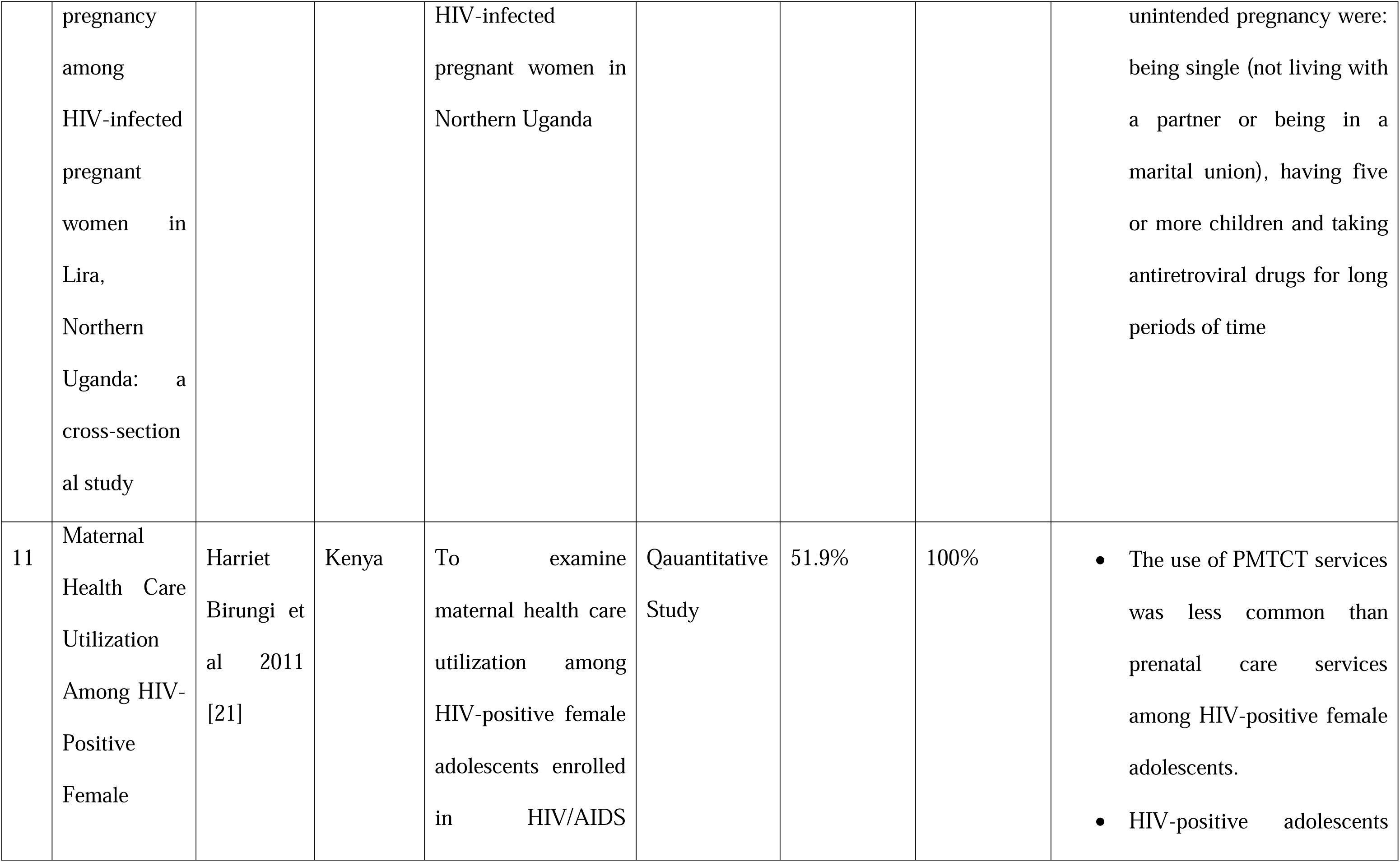
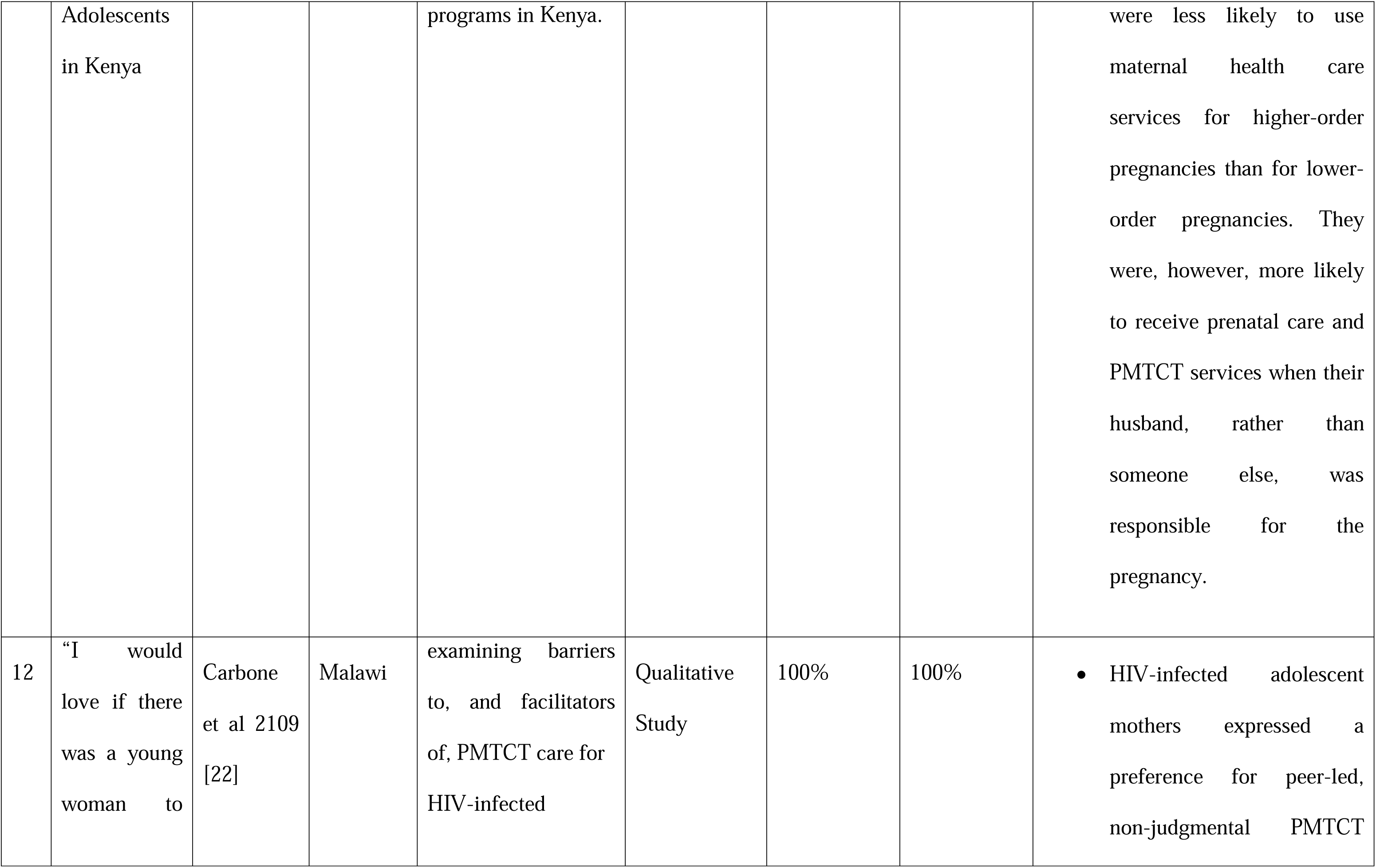
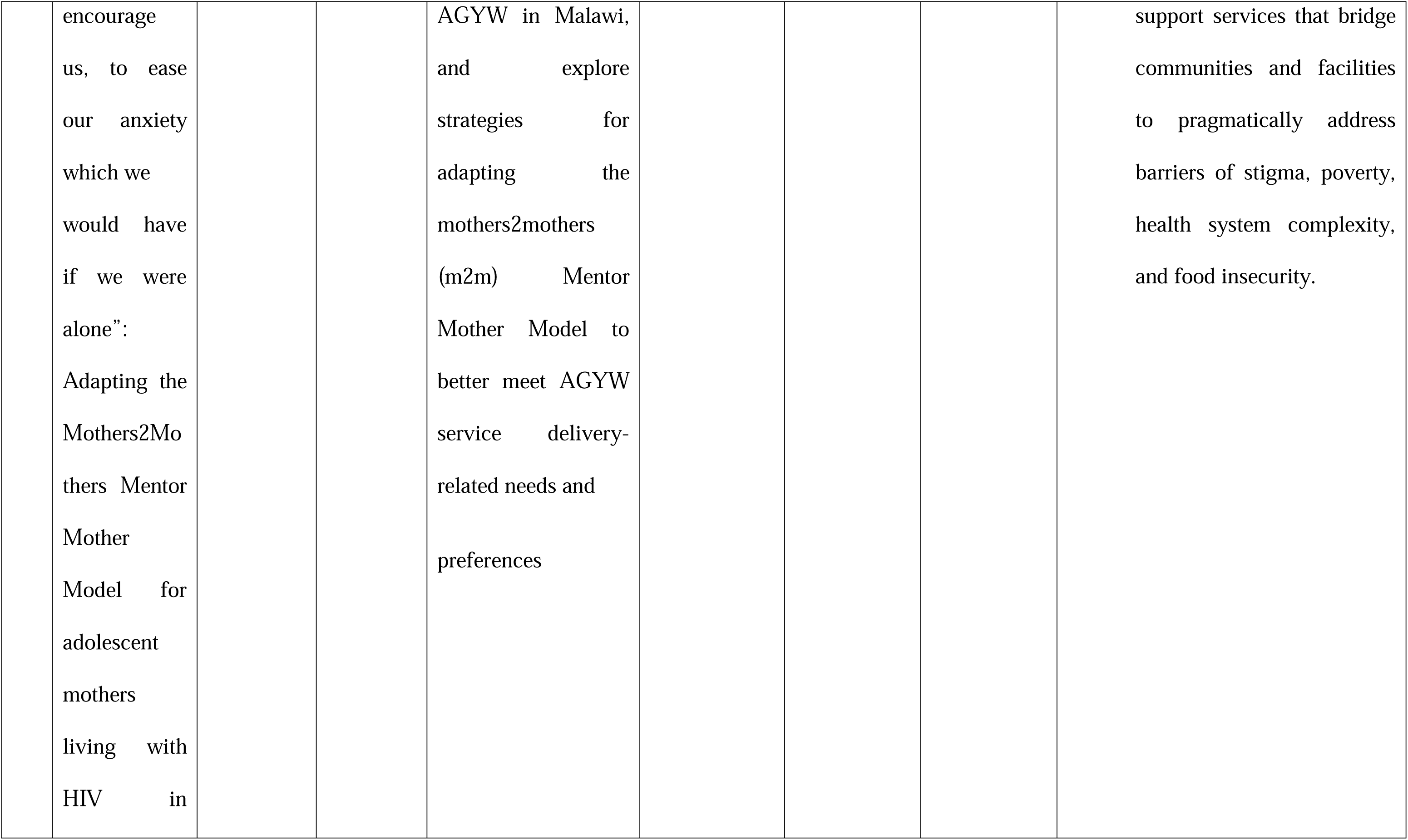
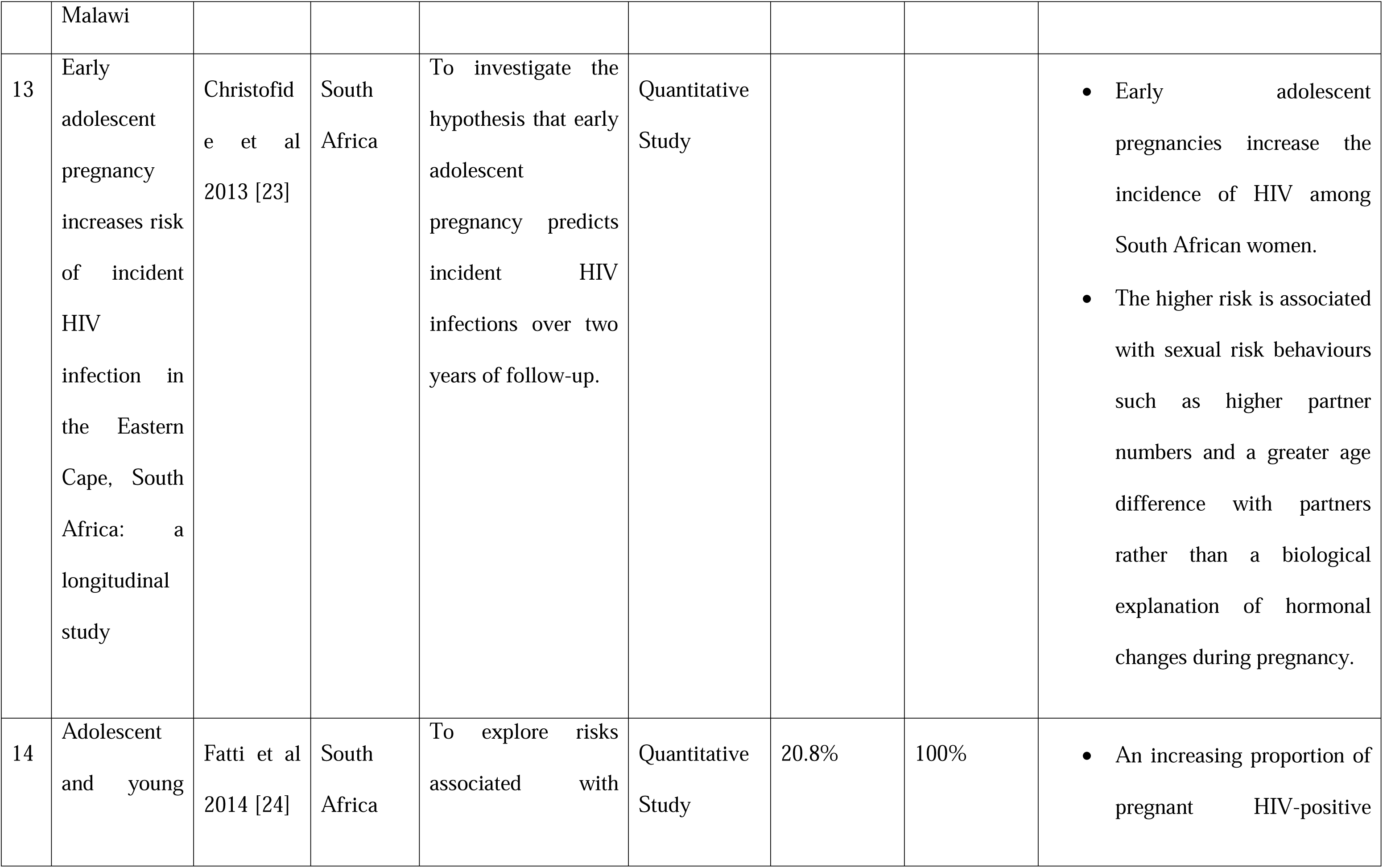
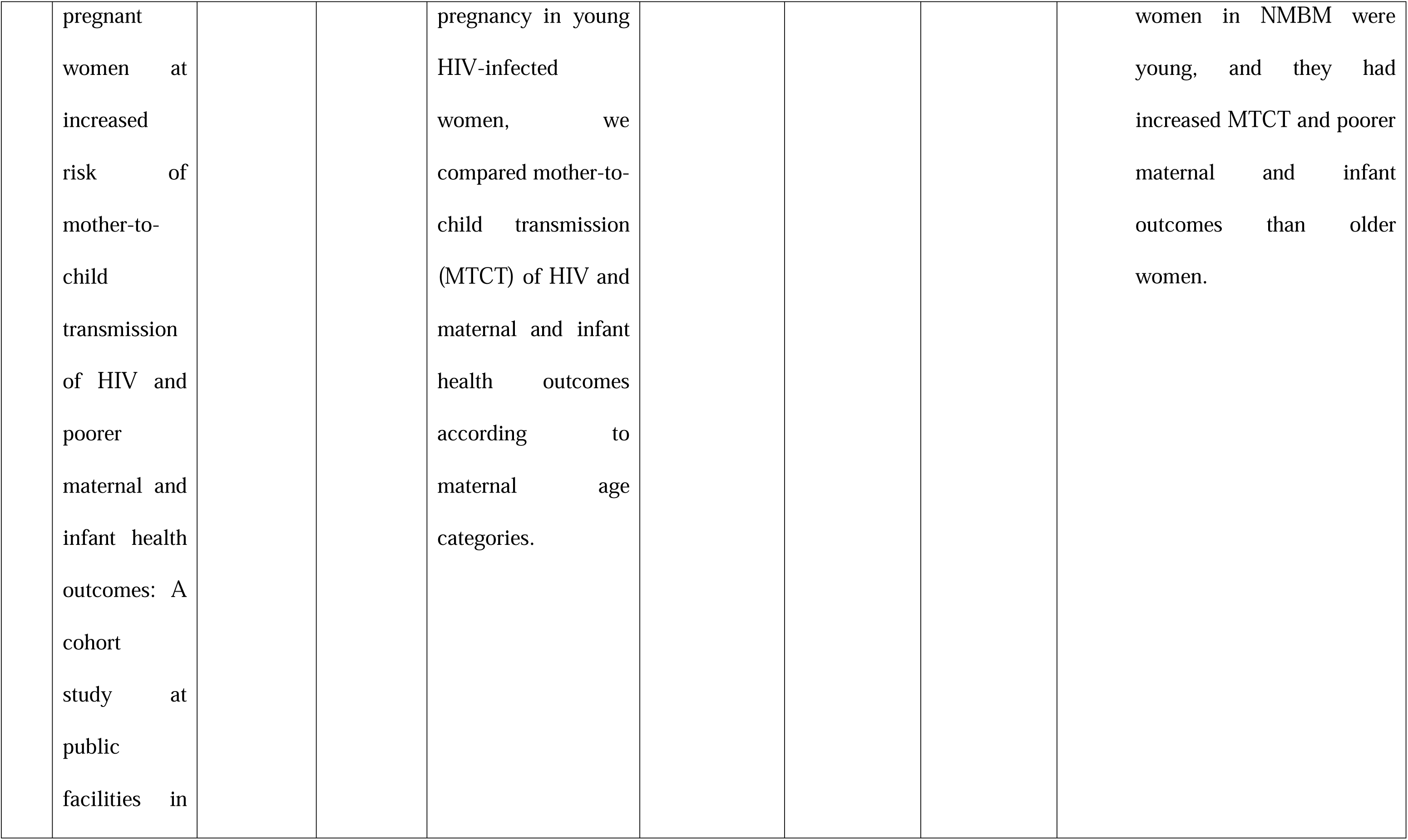
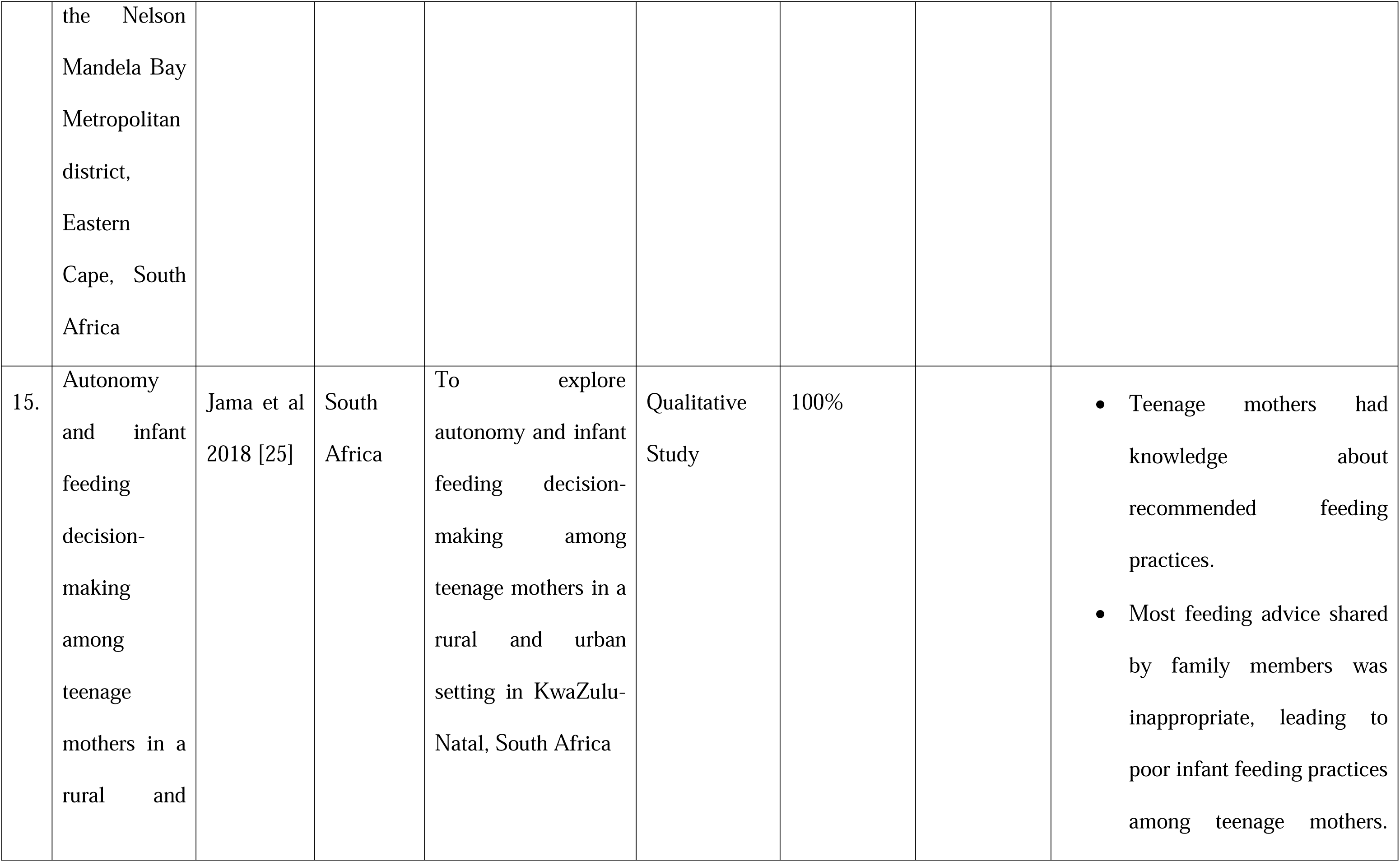
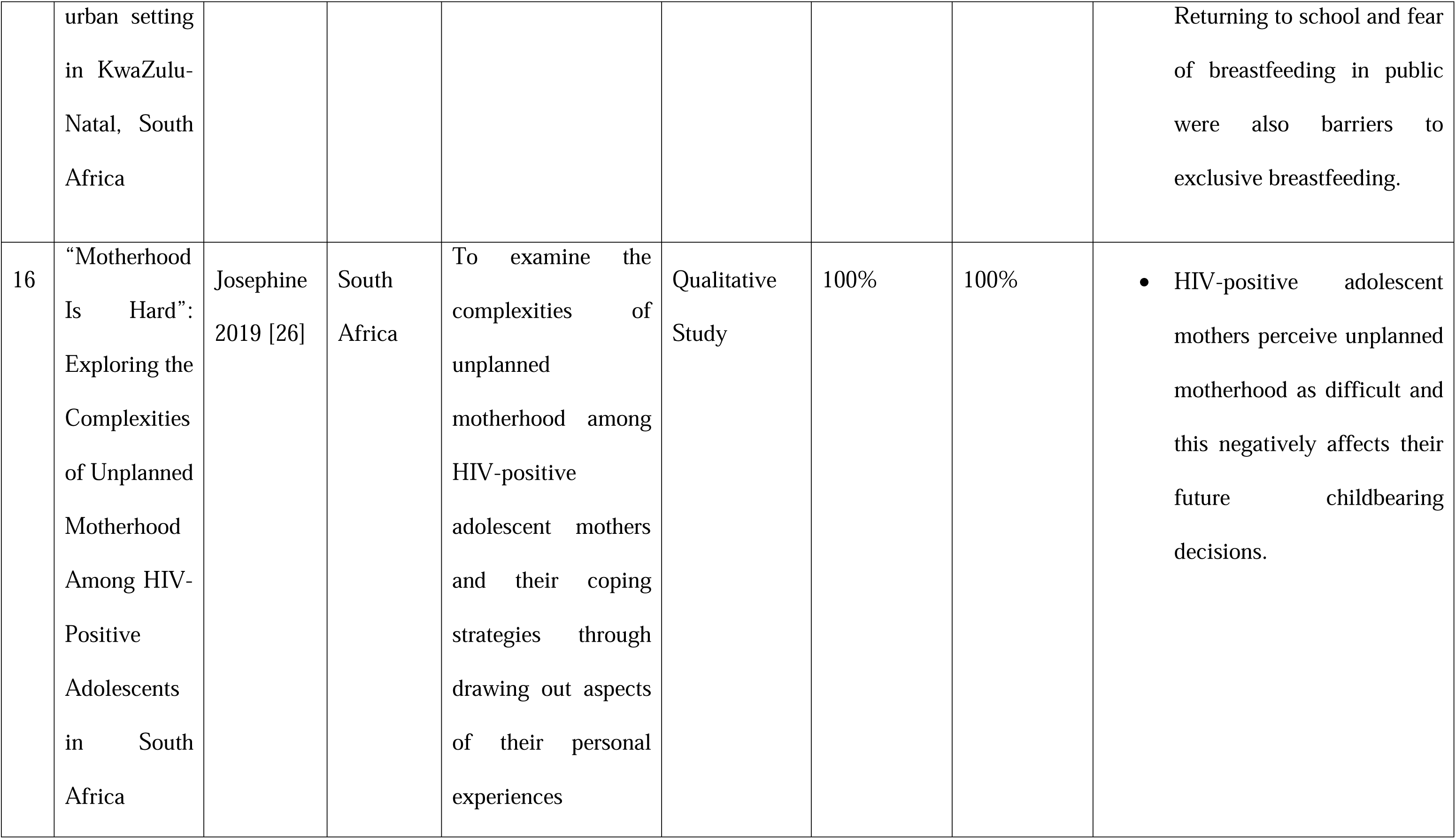
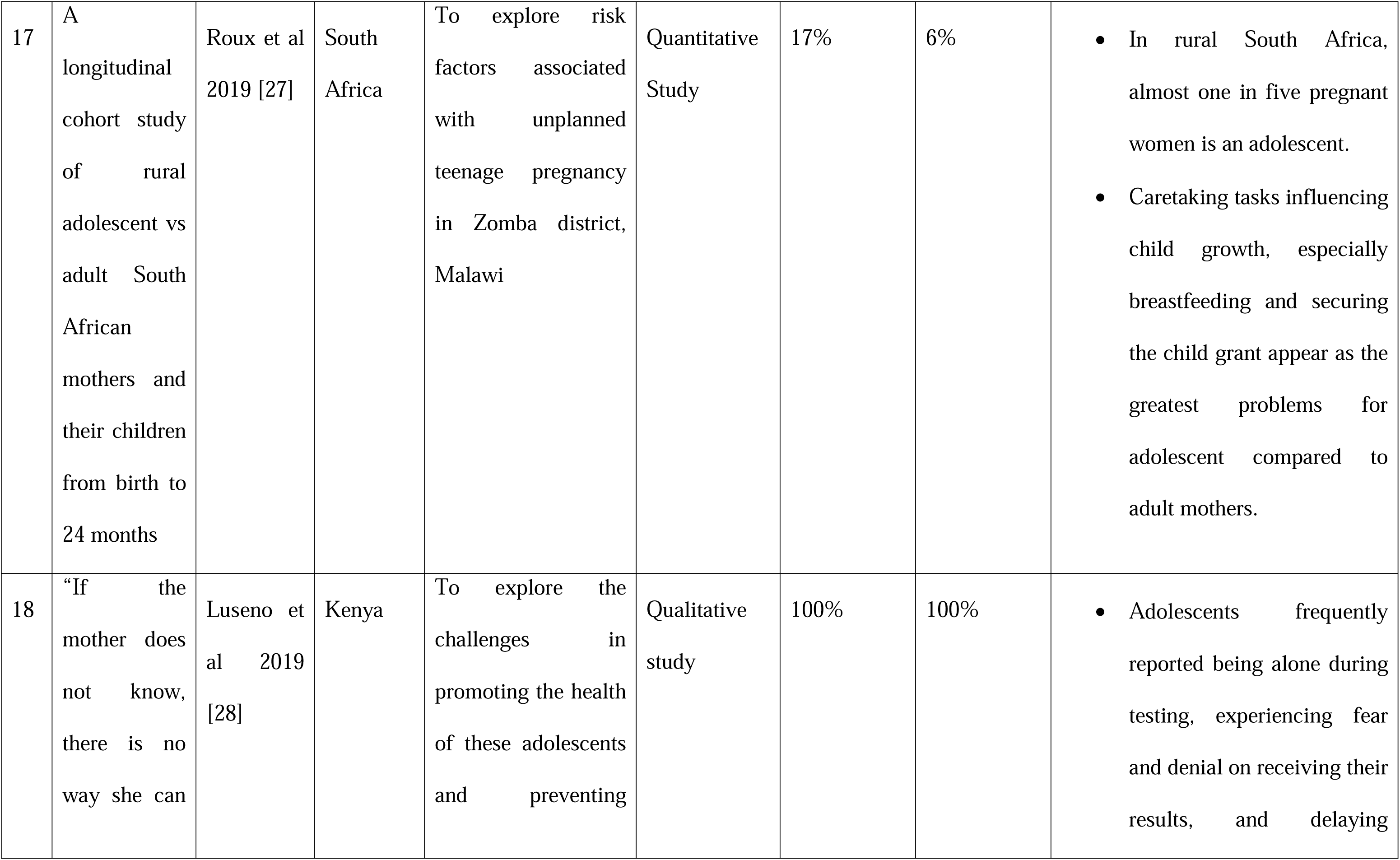
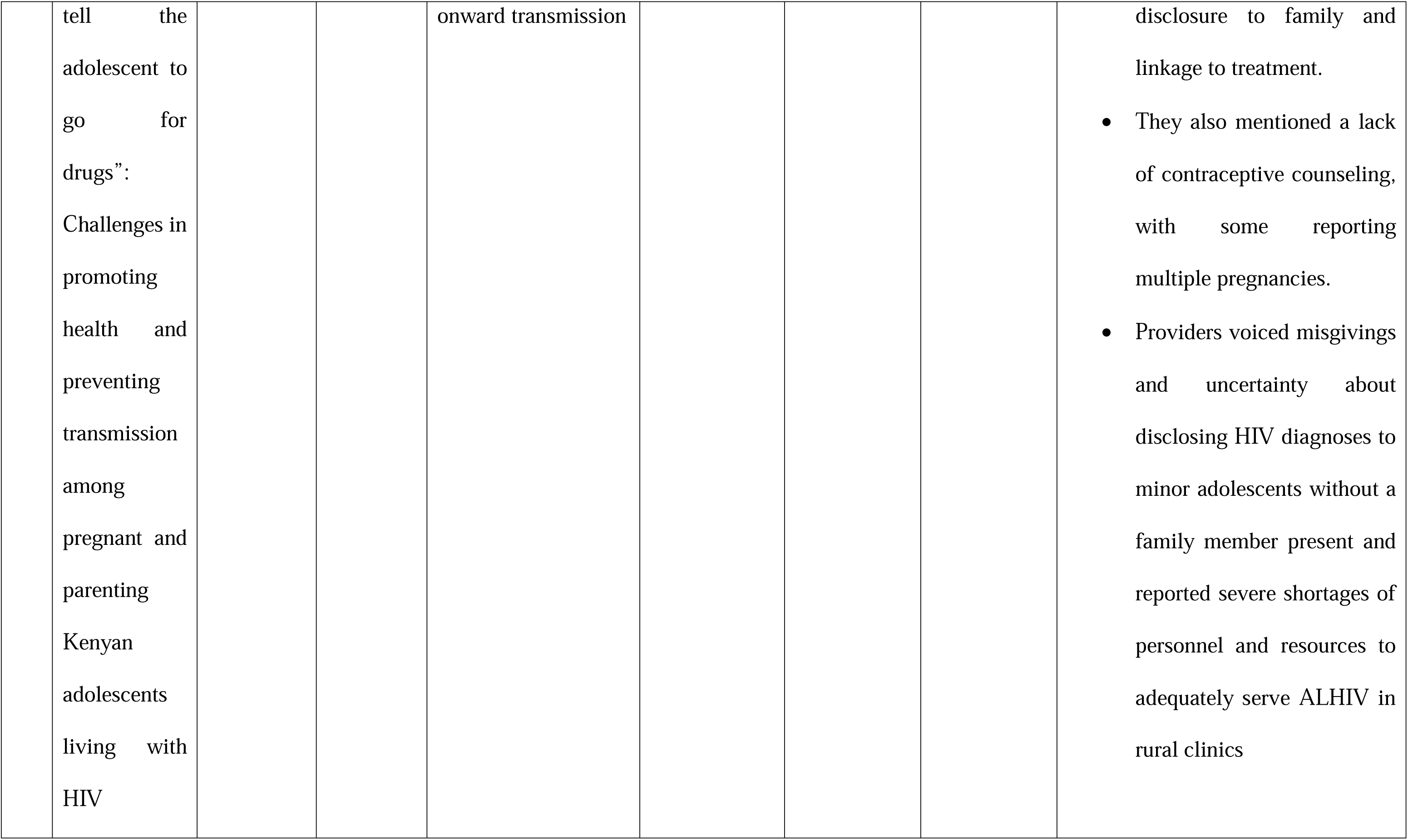
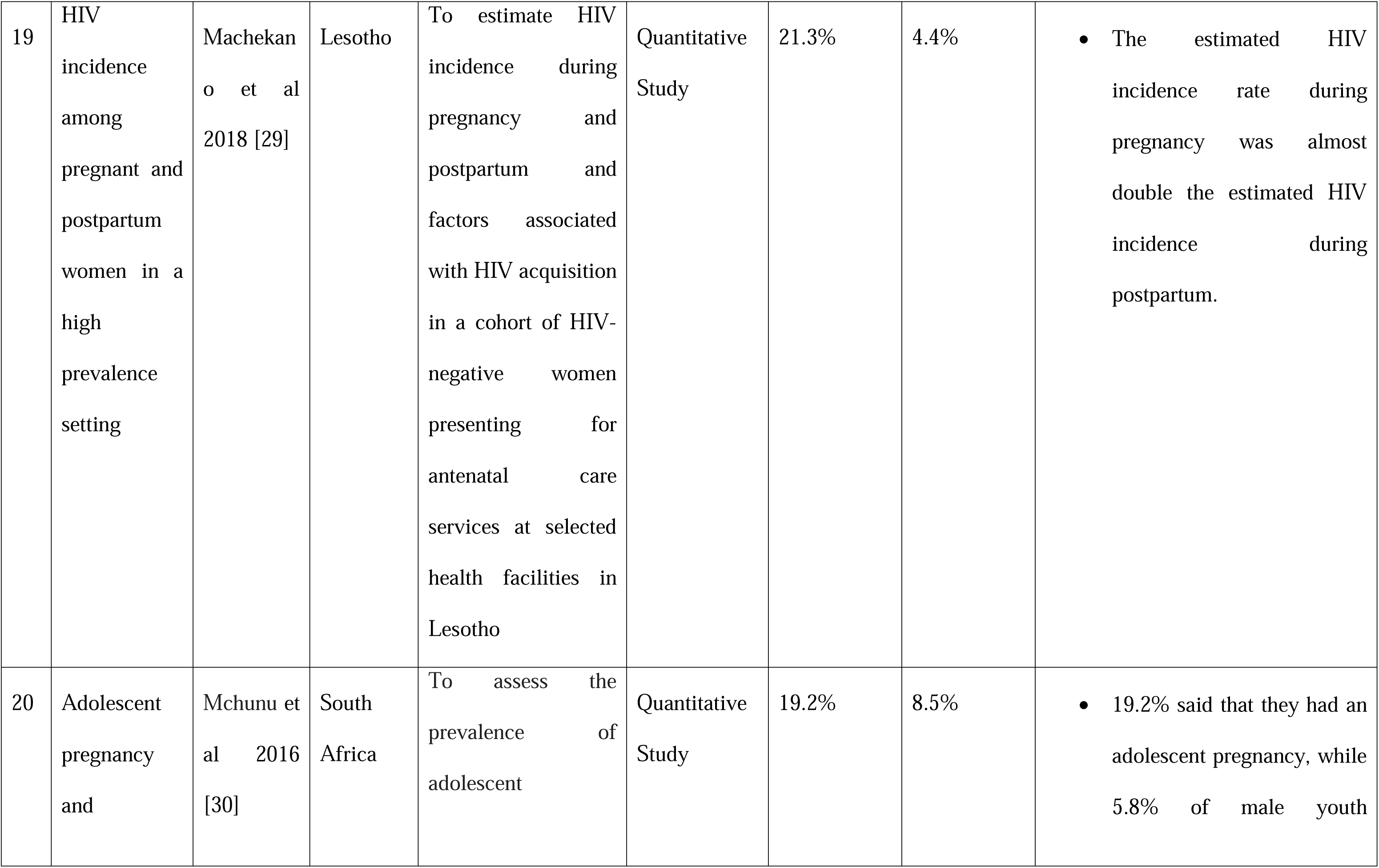
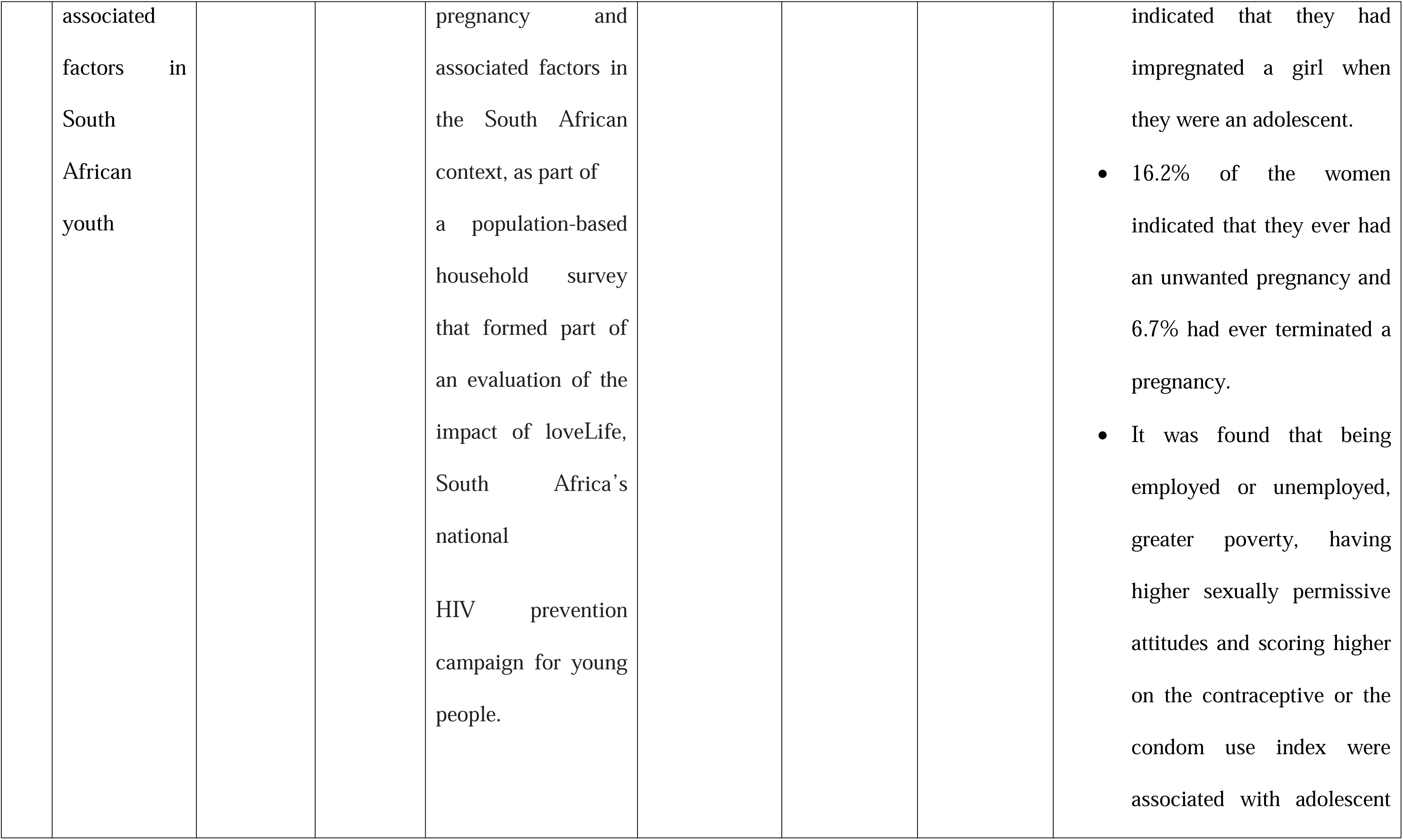
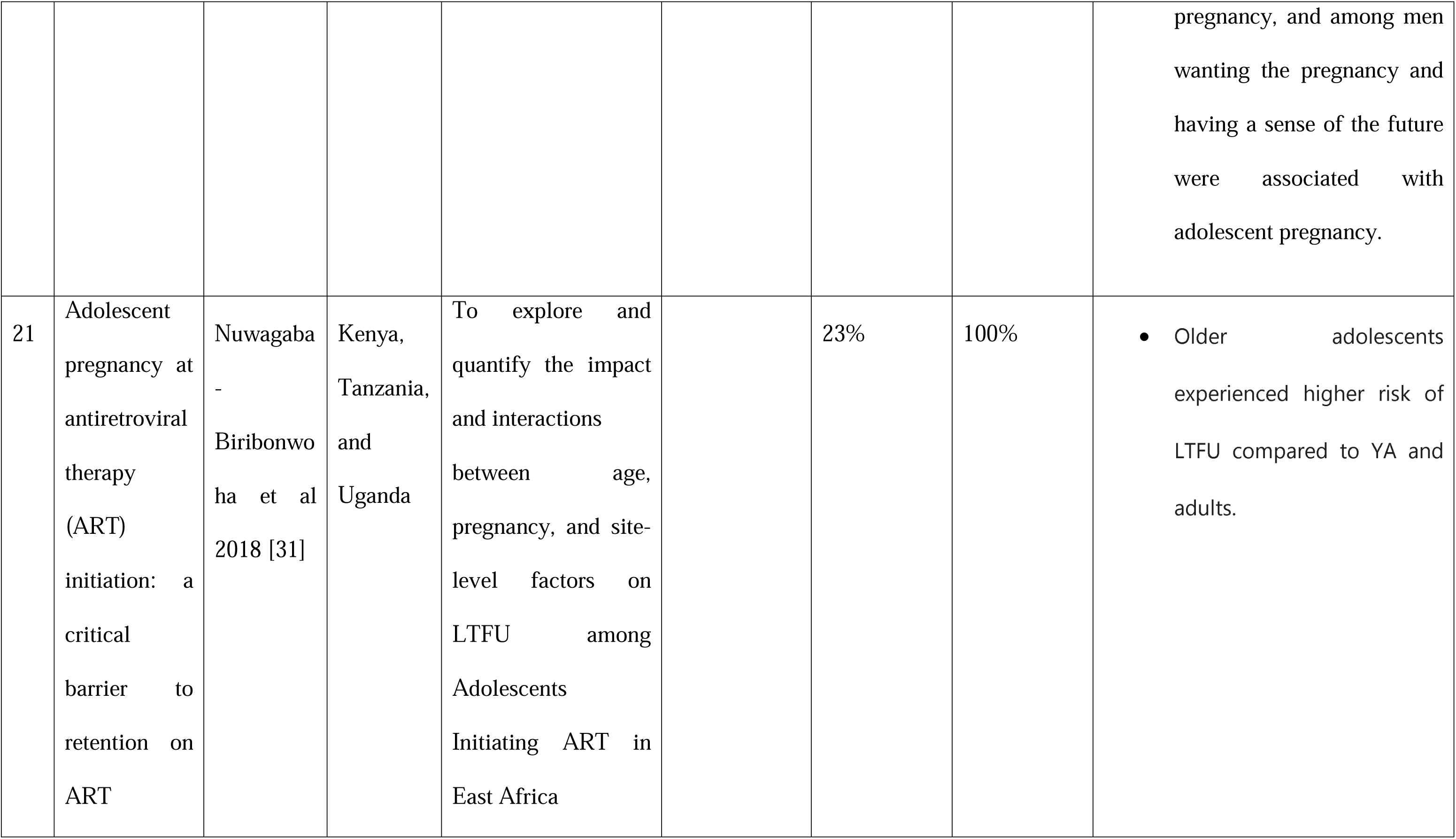
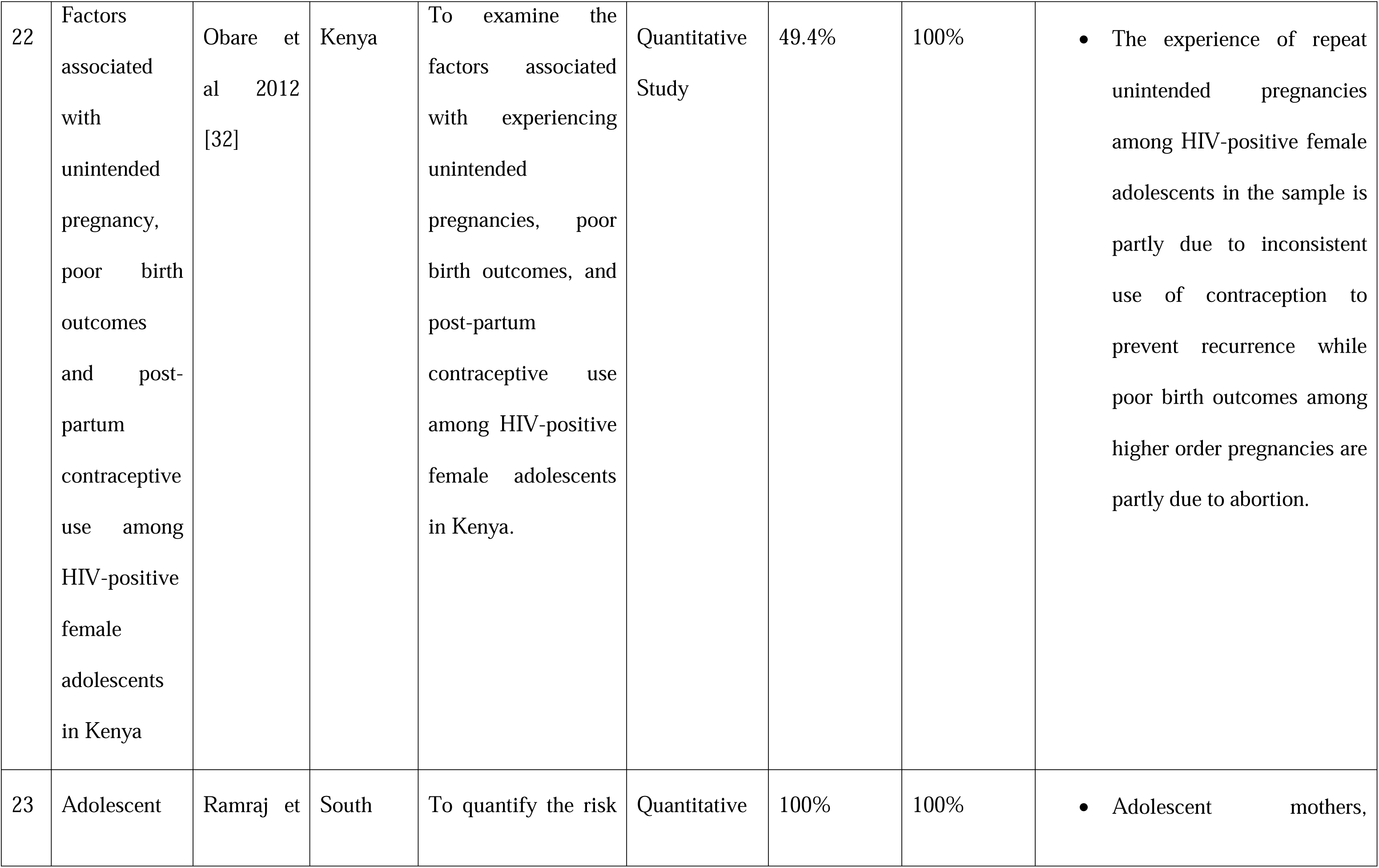
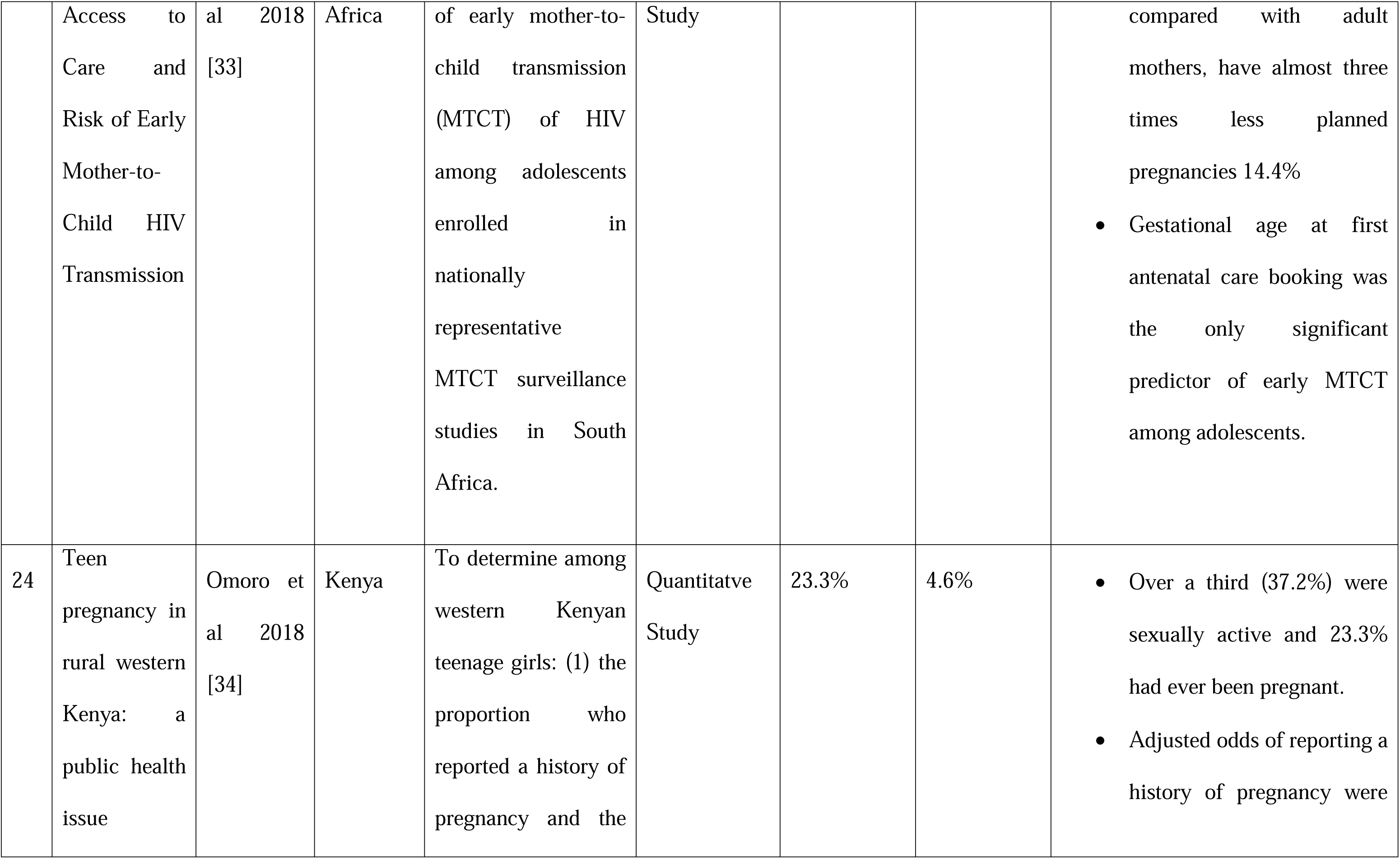
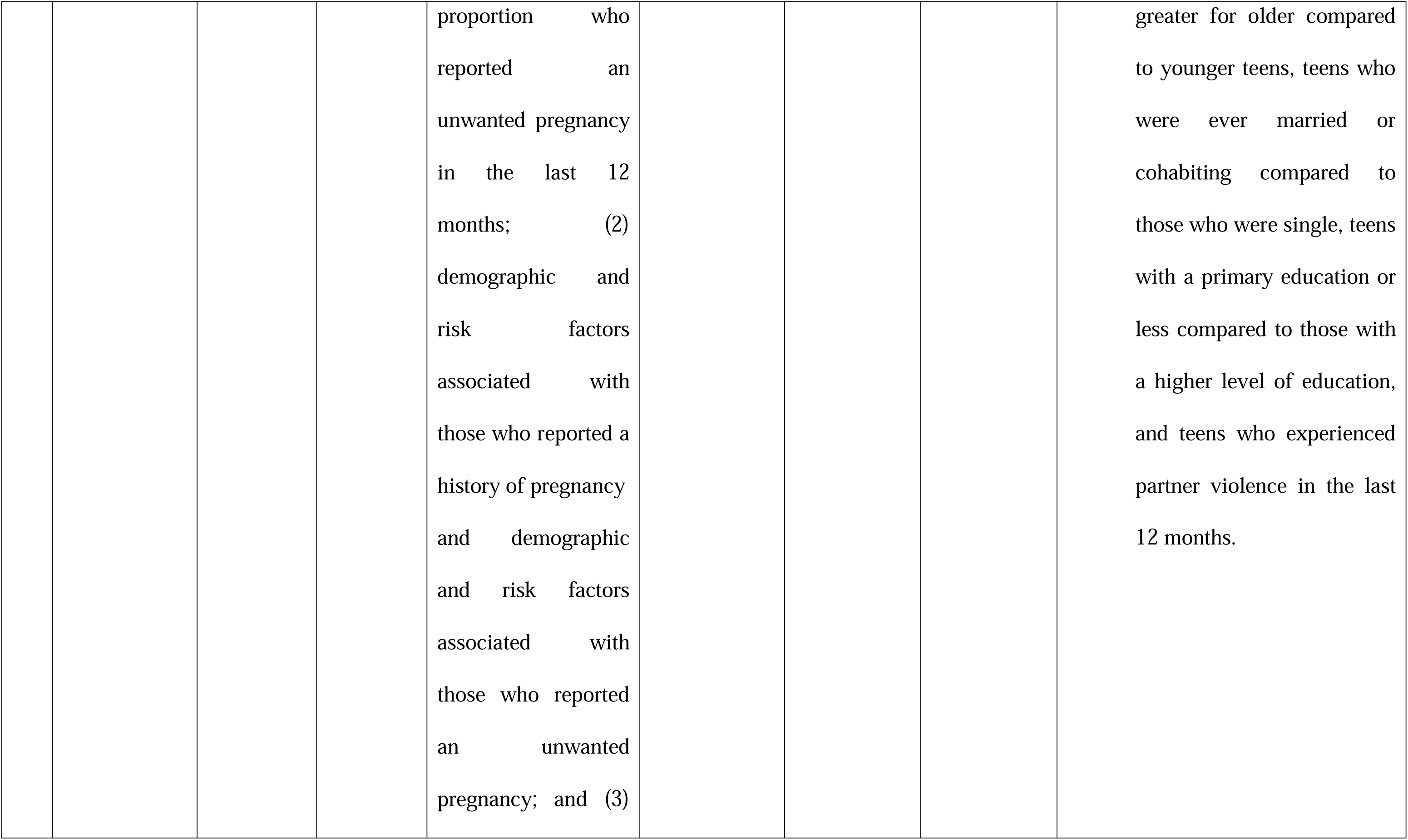
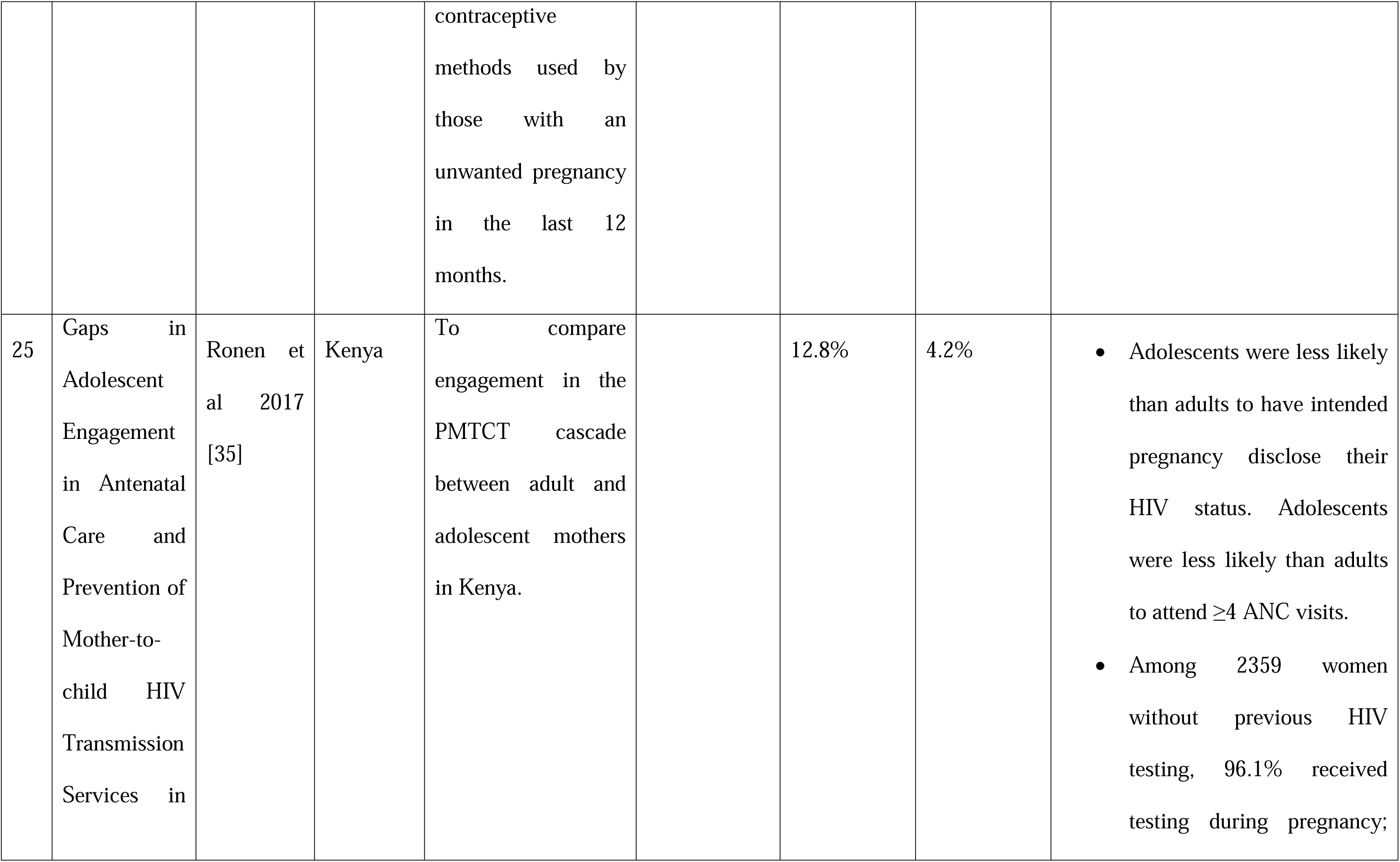
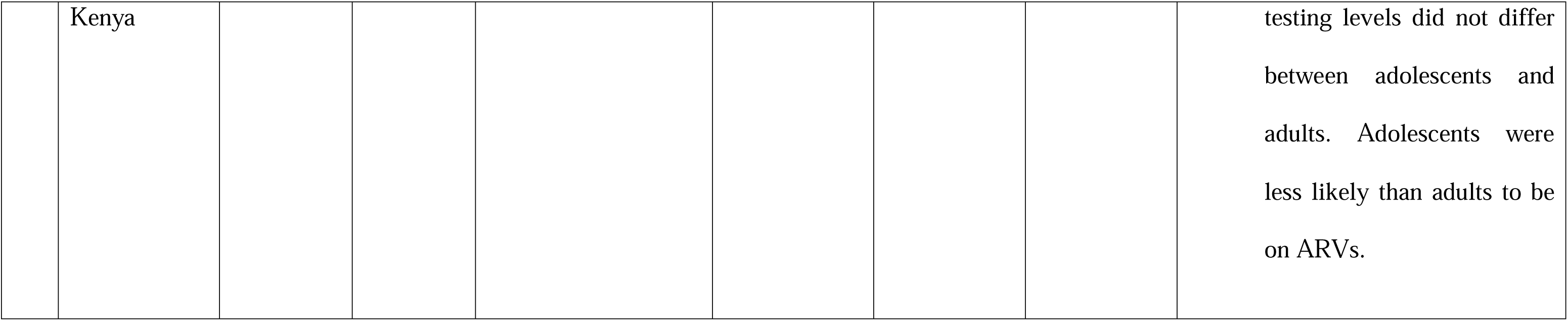
Summary of included studies.

## RESULTS

### Socio-demographic Characteristics of Participants

Figure 1 provides a Prisma Flow diagram for the study. Of the 1,560 results identified through databases (n=1,533) and hand-searching (n=27), 68 duplicates were removed, title and abstract screening for keywords relating to the topic of interest was undertaken for all articles and 52 documents were identified for full-text review. Ultimately, 27 full-text documents were excluded due to lack of relevance, a lack of data from relevant geographical locations, or not reporting on the age range of interest. In total, 25 peer-reviewed publications reported data on pregnancy among HIV-positive adolescent mothers (10–19 years) that spoke to the domains of focus (Figure 1).

### Characteristics of Included Studies

The studies that were included reported qualitative and quantitative data on the experiences of adolescent mothers affected by HIV from sub-Saharan African countries: 12 studies from South Africa [4,11,12,13,15,22,23,24,25,26,29,32], 2 from Malawi [17,21], 1 from Zimbabwe [16], 6 Kenyan studies [18,20,27,31,33,34] (n=6), 1 from Lesotho [28] 1 from Uganda [19], and 2 multi-country studies [14, 30].

The majority of the studies reported on quantitative data [4,11,12,14,15,17,18,19,20,22,24,26,28,29,31,32,33,34], five reported qualitative data [13,21,25,27,30], and one study used mixed methods [16]. The total number of participants included is 178,227, and the age range is between 12-18 years.

### Prevalence of Adolescent Pregnancies among those with HIV in Sub-Saharan Africa

Generally, the included studies consistently highlight a positive association between adolescent motherhood and HIV infection [12, 14, 22, 27]. Specifically, young women who underwent early adolescent pregnancy showed a higher likelihood of later acquiring HIV.

In a South African study carried out among 326 pregnant/parenting adolescents attending prenatal clinics and postnatal and gynecology wards, HIV prevalence was 17.8% [12]. Another study in South Africa, where 341 teenagers were interviewed, reported 82.4% having their first pregnancy and 17.6% having a repeat pregnancy. Notably, a majority of the teenagers with repeat pregnancies conceived again within 24 months of the initial or most recent pregnancy [13]. A study that tested the association between adolescent motherhood and HIV across 10 countries in Eastern and Southern Africa showed that nearly one in five (17.4%) of the adolescent girls included in the analysis were mothers and 5.8% were adolescent mothers with HIV [15].

In South Africa, the prevalence of pregnant adolescent girls who are living with HIV increased every year. In 2016, 38.8% of pregnancies occurred in adolescents ≤16 years, while 39.1% were within the 17-19 age bracket [16]. A comparable trend was observed in Zimbabwe, where 33% of young people living with HIV, aged 15 to 24, had experienced pregnancy [17]. Conversely, a study in Malawi focused on female sex workers (FSWs) revealed that 14.7% of them were between the ages of 13–19 and were living with HIV. Additionally, among the 363 participants, 52.3% had encountered their first pregnancy before turning 19 years old [18].

In studies carried out in Kenya, the highest prevalence of pregnancy in adolescents living with HIV was observed between the ages of 17 to 19 years [19,33]. Other studies conducted within the country reported that the majority of the pregnancies were reported by adolescents aged 18 or older, although most of the pregnancies had occurred when the respondents were 17 years and below [21,32]. Similarly, a longitudinal study carried out in South Africa over two years of follow-up reported that two hundred and fifty-one young women (17.74%) experienced a pregnancy when they were under the age of 19 [23].

Within Uganda, a study involving 518 HIV-positive pregnant women revealed that 6.8% were under the age of 20 [20]. In Kenya, a sampling of 498 HIV-positive women from both primary national and secondary Nyanza oversample surveys found that 21 individuals (4.2%) were adolescents [34].

Socioeconomic factors were also found to be associated with adolescent pregnancy. Low-income earners, those with no income, and those with lower levels of education were more likely to have early pregnancy in comparison to their counterparts [12,16,20,29,33,34]. They also experienced more physical and sexual violence [22, 26,33].

Moreover, the impact of early childbirth extended to subsequent pregnancies among adolescents living with HIV. Studies indicated that having children at a young age significantly influenced the likelihood of repeat pregnancies among this demographic [11, 12]. As the age of adolescents increased, so did the odds of experiencing repeated pregnancies, suggesting a correlation between age and the occurrence of repeat pregnancies.

### Impact of Pregnancies on Clinical Outcomes and Well-being

Poor mental health has been found to be higher among adolescent mothers. Probable common mental disorder was higher among adolescent mothers living with HIV (23.0%) in comparison to non-mothers and mothers who were not living with HIV. Adolescent mothers were shown to have higher prevalence of probable mental health comorbidities regardless of their HIV status, indicating that the experience of pregnancy and motherhood may contribute to a heavier burden on their mental health [4].

Qualitative data revealed that adolescent mothers living with HIV often cannot remain at home with their parents, either for reasons of overcrowding in the home, abuse, neglect, or financial difficulties. They are often averse to pregnancies because it bring emotional and physical burden to the already burdened person [14, 17]. Adolescent mothers’ safety needs are not met. Some of them have had to stay on their own with their babies or move to unsafe communities because of the threat of violent acts against them [14].

Reports from included studies show that HIV-infected pregnant adolescents have challenges with long-term contraceptive continuation and consequently poorer prevention of mother-to-child HIV transmission (PMTCT) service outcomes and higher rates of mother-to-child HIV transmission among infants compared to HIV-infected pregnant adults, likely due to the poor knowledge on care among them [13, 15, 16, 19]. Adolescent mothers have lower adherence to ANC visits and ART treatments, [16,21,31, 34, 35]. This is corroborated by another study which reported a significant number of pregnant adolescents with HIV had severe immunosuppression (CD4 count 200 cells/L) or increased VLs (400 copies/mL) [15]. Adolescents also had increased risks of maternal mortality, first presentation in labour, and stillbirth. [23,31]

Seven studies reported the low use of maternal health care services, including use of PMTCT, skilled attendance and postnatal/postabortion care for pregnancies which might end in adverse outcomes such as miscarriage, stillbirth or abortion among HIV-positive female adolescents [12, 14, 15, 18, 20, 30, 32, 34].

## DISCUSSION

The scoping review has presented an extensive synopsis of all available literature on adolescent mothers living with HIV in sub-Saharan Africa through the prevalence of adolescent pregnancies, including the risk factors, impacts on clinical outcomes and well-being, and the availability and effectiveness of reproductive health services.

The prevalence of pregnancies among adolescents living with HIV in Africa has long been a recognized concern. While prior studies have identified the co-occurrence of adolescent pregnancies and HIV, this review delves deeper into the dynamics of prevalence. For example, included studies reported the ages of 17 to 19 years to have the highest prevalence rates of adolescent pregnancy [18,34], while others reported that the majority of the pregnancies had occurred when the respondents were aged 17 years and below [20,22,31]. These disparities could be attributed to differences in healthcare infrastructure, HIV testing, and antiretroviral therapy availability [36,37].

This study sheds light on a significant disparity in the representation of adolescents within certain demographics, particularly in studies involving pregnant individuals living with HIV in South Africa. In both cases cited, adolescents comprised a notably lower percentage compared to their older counterparts. In the first instance, among pregnant adolescents and young pregnant women living with HIV, only 20.8% fell within the adolescent age range (13-19 years), while the older cohort (20-24 years) represented a higher proportion at 32.6%. This disparity indicates a potential discrepancy in the impact of HIV within different age groups, suggesting varying levels of vulnerability or access to healthcare services between adolescents and slightly older women.

Similarly, in the longitudinal study that assessed the challenges faced by adolescent versus adult mothers in rural South Africa during the initial 24 months post-birth, adolescents accounted for only 17% of the mothers. This percentage was notably lower when considering HIV seropositive cases, where only 6% of adolescent mothers were identified as HIV positive. This finding, in contrast to their adult counterparts, highlights a significant difference in HIV prevalence between adolescent and adult mothers. The lower representation of adolescents in these studies might reflect several underlying factors. There could be differences in healthcare-seeking behaviors, access to reproductive health education, or social support systems between adolescents and older individuals. It could also signify varying levels of vulnerability or risk perception among these age groups regarding HIV and pregnancy.

Understanding these disparities is crucial for developing targeted interventions and support systems that address the specific needs and challenges faced by adolescents, especially pregnant adolescents living with HIV. It emphasizes the necessity of tailored healthcare services and comprehensive support frameworks to bridge the gap and ensure equitable care and resources for all demographics, irrespective of age.

The study also showed that HIV prevalence was higher among adolescent mothers in comparison to adolescent non-mothers. This is in contrast to a study in South Africa where adolescent mothers were less likely to be living with HIV compared to never-pregnant adolescents within the sample (55.5% vs. 73.3%). The prevalence of adolescent motherhood in the sample was 15.2%, and almost all (98.8%) pregnancies were unplanned or unwanted [4]. Studies have shown that adolescents were more likely to report having had at least one unwanted pregnancy [17, 29].

Existing studies have shown that early sexual debut is a significant risk factor for both unintended pregnancies and HIV infection due to various factors such as limited knowledge about safe sex practices, lack of access to contraceptives, and potential engagement in risky sexual behaviours [38,39]. This may also be linked to adolescent mothers learning of their HIV status for the first time when they confirm that they are pregnant [30,40]. This review also highlights that adolescent mothers are more likely to be involved in relationships where they have poor relationship power, gender inequitable norms, intimate partner violence, and little HIV preventive communication with higher-risk partners [43,44, 41,42].

The association between lower education levels and higher repeat pregnancies among adolescents living with HIV, suggests that improving educational opportunities and knowledge about reproductive health may play a crucial role in preventing repeated pregnancies in this vulnerable group [45,46]. Additionally, the role of poverty in early pregnancies is a common thread in many studies. Poverty often limits access to contraceptives, healthcare, and comprehensive reproductive health education [46,47]. Targeted interventions that address economic disparities are imperative. The prevention of adolescent pregnancies and HIV goes beyond healthcare; it involves broader socio-economic and educational support. The findings of this review emphasize the importance of holistic interventions that encompass not only clinical care but also social and economic empowerment

Mental health challenges among adolescent mothers living with HIV are a shared concern, as evidenced by the high prevalence of probable mental health disorders in this population [4,48]. Stigma, lack of social support, poverty, and the timing of HIV diagnosis may contribute to these mental health challenges [48].

Hindrances in relation to access, utilization, and the quality of services are common themes in this review. Issues such as service availability in rural areas and shortages of well-trained personnel are shared concerns. Geographic disparities and socio-economic barriers also hinder access to essential care [13,27]. While the South African government provides free services such as counselling, child support grants, food parcels, housing and social grants to adolescent mothers living with HIV, these services are often not accessible to those who stay in rural communities and are far from social welfare offices [13]. Smaller rural clinics, which are often more accessible to these adolescents, also have to deal with the shortage of well-trained personnel and insufficient staffing as a hindrance to achieving effective adolescent care [27].

The included studies underscore notable negligence in meeting the reproductive health needs of individuals transitioning from childhood to adulthood. Often, practical solutions for sexually active adolescents, or those intending to be sexually active, are lacking. Emphasis tends to revolve around advocating abstinence from sexual intercourse without providing comprehensive and practical guidance or support [30, 31].

## RECOMMENDATIONS

This study suggests that larger facilities, such as referral hospitals, may be better equipped to offer adolescent-friendly PMTCT services that emphasize confidentiality, address stigma related to HIV infection and adolescent pregnancy, and integrate models of increasing social support. Contact with antenatal and postnatal care services and HIV care services for adolescents living with HIV also provide an opportunity to merge differing branches of health service through providing screening, monitoring, and referral for mental health.

The results from this study also recommend the need for integrated mental health services within reproductive health care programs tailored to the unique needs of adolescent mothers in SSA. Challenges faced by adolescent mothers living with HIV, including accelerated disease progression and heightened mental health issues, were highlighted in this review. Research has repeatedly identified stigma, lack of social support, parenting stress, and knowledge gaps as factors contributing to mental health challenges [4,48]. This highlights the necessity for a more holistic approach, one that goes beyond advocating abstinence and includes comprehensive reproductive health education, access to contraceptives, support for safer sexual practices, and tailored interventions for adolescents living with HIV who are navigating parenthood. Addressing these gaps is vital to ensure the well-being and reproductive health empowerment of adolescent mothers living with HIV.

## Data Availability

The authors confirm that the data supporting the findings of this study are available within the article and its supplementary materials.

## Acknowledgement

This work was supported by the Nigerian Institute of Medical Research, Lead City Univeristy, and Fogarty-IeDEA Mentorship Program (FIMP), West Africa.

## Data Availability Statement

The data used to support the findings of this study are available from the corresponding author upon request.

## Appendix: Search Terms

### PubMed-59

|  | Keywords | Search Strings |
| --- | --- | --- |
| 1 | Prevalence | ((TS=(Prevalence )) OR TS=(Epidemiology)) OR<br>TS=(Frequency) and Preprint Citation Index (Exclude –<br>Database) |
| 2 | Adolescent pregnancy | ((((((((((((TS=(Adolescence, Pregnancy )) OR TS=(Teen<br>Pregnancy)) OR TS=(Pregnancy, Teen)) OR<br>TS=(Adolescent Pregnancy)) OR TS=(Pregnancy,<br>Adolescent)) OR TS=(Pregnancy, Teenage)) OR<br>TS=(Teenage Pregnancies)) OR TS=(Preteen Pregnancy))<br>OR TS=(Pregnancy, Preteen)) OR TS=(Child Pregnancy))<br>OR TS=(Girls pregnancy)) OR TS=(Adolescent mothers))<br>OR TS=(Pregnancy unplanned)) OR TS=(Pregnancy<br>unwanted) and Preprint Citation Index (Exclude –<br>Database) |
| 3 | HIV | ((((((((((((TS=(Human Immunodeficiency Virus)) OR<br>TS=(Immunodeficiency Virus, Human)) OR<br>TS=(Immunodeficiency Viruses, Human)) OR TS=(Virus,<br>Human Immunodeficiency)) OR TS=(Viruses, Human<br>Immunodeficiency)) OR TS=(Human Immunodeficiency |
|  |  | Viruses)) OR TS=(Human T Cell Lymphotropic Virus Type III)) OR TS=(LAV-HTLV-III)) OR TS=(Lymphadenopathy-Associated Virus)) OR TS=(Human T Lymphotropic Virus Type III)) OR TS=(AIDS Virus)) OR TS=(Acquired Immune Deficiency Syndrome Virus)) OR TS=(HTLV-III) and Preprint Citation Index (Exclude – Database) |

### CINAHL= 0

|  | Keywords | Search Strings |
| --- | --- | --- |
| 1 | Prevalence | TI ( prevalence or incidence or epidemiology or frequency or occurrence or statistics ) OR TI prevalence rates |
| 2 | Adolescent Pregnancy | TI adolescent pregnancy or teenage pregnancy or teen pregnancy |
| 3 | HIV | TI HIV or aids or acquired human immunodeficiency syndrome or human immunodeficiency virus |

### WEB OF SCIENCE

#### Web of science (Number)

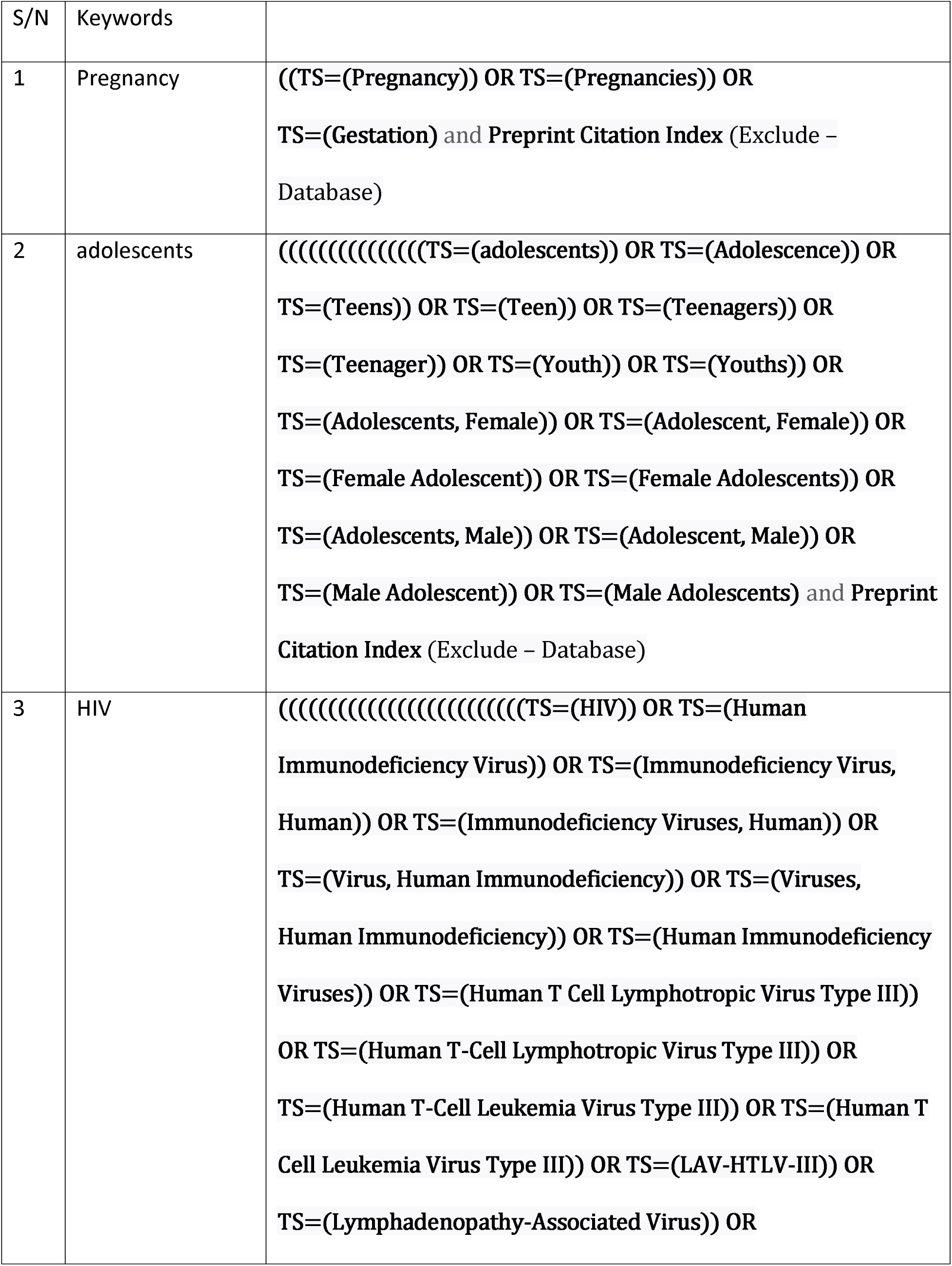
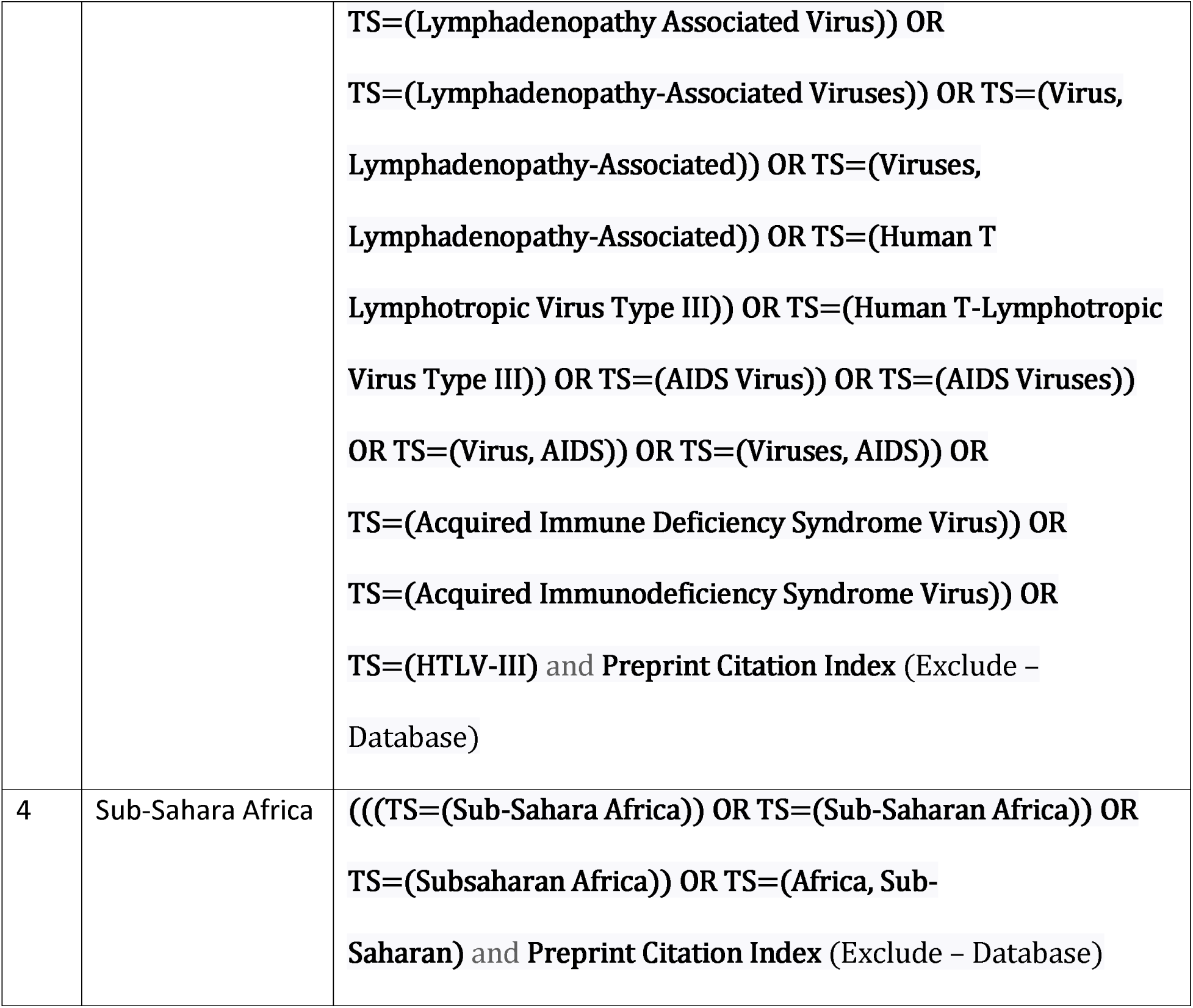

### SCOPUS = 462

(TITLE-ABS-KEY(prevalence) AND TITLE-ABS-KEY(adolescent) AND TITLE-ABS-KEY(Pregnancy) AND TITLE-ABS-KEY(HIV) AND TITLE-ABS-KEY(Africa)) AND PUBYEAR > 2001 AND PUBYEAR < 2023 AND (LIMIT-TO ( DOCTYPE,“ar”)

